# Avoidable childhood respiratory-infection deaths: a frontier analysis of episode-fatality ratios in 204 countries, 1990–2023

**DOI:** 10.64898/2026.09.01.26361882

**Authors:** Deze Li, Hao Chen, Xiaotong Wang, Chen Shen

**Author notes:** Correspondence to: Chen Shen, Beijing Pediatric Research Institute, Beijing Children’s Hospital, Capital Medical University, Beijing, China.

## Abstract

**Background:** Respiratory infections remain the leading infectious cause of death among children and adolescents, yet the share of these deaths that could be averted with currently feasible care is not routinely quantified. Existing amenable-mortality frameworks rely on cause lists and population-level mortality benchmarks and do not exploit information on how many episodes occur. We propose an episode-fatality-ratio (EFR) frontier approach and apply it to lower respiratory infections (LRI), whooping cough (pertussis) and upper respiratory infections (URI) in 204 countries, 1990–2023.

**Methods:** For each cause, country and year we computed EFR = deaths/incident episodes using Global Burden of Disease (GBD) 2023 estimates for ages 0–19 years. The frontier was defined as the 10th-percentile country EFR within each GBD super-region, cause and year; avoidable deaths = max(0, deaths − episodes × frontier EFR). Primary estimates are deterministic; 95% uncertainty intervals (UIs) come from 2,000 Monte Carlo draws. Sensitivity analyses varied the frontier percentile, applied an aspirational global frontier, constructed pertussis counterfactuals, and recomputed all estimates within the single under-5 age band.

**Findings:** In 2023, 333,803 childhood deaths from lower respiratory infections (95% UI 289,123–417,460; 46.9% of LRI deaths) were avoidable. Summing the three causes deterministically gives 391,034 avoidable deaths (46.5% of 840,444); the combined figure is a deterministic sum, and a UI is available for the LRI component only. The pertussis (43,958; 39.0%) and URI (13,273; 81.0%) estimates are secondary: their deterministic point values fall below their own Monte Carlo intervals and the underlying death estimates carry very wide uncertainty (global pertussis UI 12,545–321,874). Avoidable deaths fell from 1,050,468 (44.9%) in 1990, but between 2019 and 2023 the avoidable share for LRI+URI barely moved (48.7% to 47.7%) while absolute avoidable deaths fell 14.5%, a pattern consistent with stalled convergence to the frontier. Sub-Saharan Africa plus South Asia held 73.1% of avoidable deaths in 2023 versus 41zzz

## Introduction

Lower respiratory infections (LRI) remain the leading infectious cause of death in children worldwide. Global Burden of Disease (GBD) 2023 estimates attribute 711,228 deaths (95% uncertainty interval 507,283–958,451) among 0–19-year-olds to LRI in 2023, down from 2,035,714 in 1990, a 65% decline driven jointly by falling incidence and improving survival once infected.^1,2^ Pertussis, a vaccine-preventable respiratory infection against which routine immunization has been in place for six decades,^3,4^ still caused an estimated 112,839 deaths in this age group in 2023,^1^ while upper respiratory infections (URI) are nearly ubiquitous (5.6 billion episodes annually) yet rarely fatal.^1^ The unfinished communicable-disease agenda among children and adolescents has been well documented,^5^ but how much of the residual respiratory-infection mortality could realistically be averted with care that is already demonstrably achievable remains less precisely quantified.

The concept of deaths that need not occur given available medical knowledge dates to Rutstein and colleagues’ 1976 proposal of “untimely and unnecessary deaths” as sentinel events of health-system performance, in which a death from a listed condition signals a possible failure of prevention or care warranting investigation.^6^ Nolte and McKee developed this tradition into the amenable-mortality framework that underpins contemporary global health measurement:^7^ the Lancet Global Health Commission on High Quality Health Systems estimated that 8.6 million deaths annually in low- and middle-income countries are amenable to health care, with poor-quality care now implicated in more deaths than non-utilization of services,^8^ and the GBD Healthcare Access and Quality (HAQ) Index operationalized amenable mortality as a tractable cross-country performance metric, scoring each country against the lowest observed age-specific death rates for 32 causes.^9^ Childhood respiratory infections occupy a prominent place on every such list: they are common, well understood clinically, and responsive to inexpensive interventions (antibiotics, oxygen and vaccines) that have been standard of care for decades. They therefore function as tracer conditions for the proposition that where health systems work, children should not die of pneumonia.

These frameworks, however, share two structural features that limit their resolution for respiratory infections. First, they score expert-defined cause lists against population-level mortality benchmarks, whether the fixed ceilings of the classical amenable-mortality tradition^7^ or the empirical age-specific frontier of the HAQ Index,^9^ and in either form they conflate epidemiological exposure with health-system performance. A country with many pneumonia episodes will record more “amenable” deaths than a country with few, even if both treat each episode equally well; conversely, a country that prevents episodes through vaccination or risk-factor control is rewarded twice, once in the exposure term and once in the amenable-death total. Second, the cause-list approach discards information on how many episodes actually occur. The ratio of deaths to incident episodes, the episode-fatality ratio (EFR), separates the act of falling ill from the act of dying once ill, and therefore isolates the component of mortality most plausibly responsive to case management: timely antibiotics, oxygen, supportive care and, for pertussis, vaccination preventing severe disease in the first place.^2^ In cancer epidemiology the analogous mortality-to-incidence ratio has been widely used as a proxy for survival, although its validity has been questioned on the grounds that deaths and incident cases arise from different cohorts;^10^ for acute respiratory infections, whose course from episode onset to death is measured in days to weeks, this particular objection carries little force. Incidence and mortality for LRI have both been estimated systematically within GBD,^12^ yet to our knowledge no cross-country avoidable-death accounting has exploited the episode denominator directly.

We therefore propose an EFR frontier approach to avoidable death estimation. Rather than asking whether a country’s death rate exceeds an arbitrary ceiling, we ask how many deaths would not have occurred if each country had achieved the episode-fatality ratio of the best-performing decile of countries within its own GBD super-region—an empirical, annually updatable benchmark that requires no external income or development covariates. We apply the method to three causes chosen to span the spectrum of respiratory-infection fatality: LRI as the primary high-burden cause, pertussis as a vaccine-preventable dimension, and URI as a near-zero-fatality contrast that bounds the method’s behaviour.

Three features make respiratory infections an informative test case for such a frontier. Episodes vastly outnumber deaths, so the fatality ratio is estimated against a large and comparatively stable denominator. The clinical pathway from episode to death is short and well specified, so the ratio plausibly tracks the reach and quality of case management, although it also reflects the severity mix of episodes (see Limitations). And the three causes span the full fatality spectrum, allowing the method’s behaviour to be inspected from the high-burden interior (LRI) through the vaccine-preventable margin (pertussis) to the near-zero-fatality bound (URI).

The study addresses three questions. First, how large is the avoidable burden of childhood respiratory-infection deaths in 2023 relative to empirically achieved within-region frontiers, and how has it changed since 1990? Second, where is the avoidable burden concentrated, and how has its geography shifted over three decades? Third, how sensitive are the estimates to the frontier definition, and does the residual gap reflect countries lagging behind a moving frontier or stagnation of the frontier itself? Answers to these questions bear directly on whether the remaining deaths are best addressed by extending basic interventions to lagging populations or by raising the standard of care everywhere.

## Methods

### Study design and data sources

This is a cross-sectional and trend analysis of modelled national estimates from GBD 2023,^1,11^ which synthesizes vital registration, surveillance, verbal autopsy and survey data through standardized cause-of-death and non-fatal modelling processes to produce internally consistent estimates for all locations and years. We extracted country-level deaths and incident episodes for ages 0–19 years (GBD age groups <5, 5–9, 10–14 and 15–19 years, summed with their uncertainty intervals) for three causes: lower respiratory infections (primary analysis), upper respiratory infections (low-fatality contrast) and whooping cough/pertussis (vaccine-preventable dimension). Coverage is 204 countries and territories. Anchor years are 1990, 2019 and 2023, chosen to span the pre-Millennium-Development-Goal baseline, the pre-pandemic reference year and the most recent GBD cycle, subject to data availability: LRI deaths span 204 countries for the full 1990–2023 series (a country-year file), and country-level LRI incidence is available for 1990, 2019 and 2023; URI deaths and incidence are available for 1990, 2019 and 2023; pertussis deaths and incidence for 1990 and 2023. Wherever results combine causes or years with different coverage, the scope is stated explicitly alongside the estimate. All inputs are publicly released modelled aggregates; no individual-level data were used and ethics review was not required.

### Episode-fatality ratio

For each cause c, country i and year y, EFR = deaths/incident episodes. The EFR is not a clinical case-fatality ratio. GBD incident-episode denominators include mild community and outpatient episodes (81.0 million LRI episodes and 5.61 billion URI episodes globally in 2023), so EFRs lie far below facility-based case-fatality ratios and the two quantities are not interchangeable.^11^ The EFR instead summarizes, for an entire national health system and catchment population, the probability that an episode of illness ends in death.

### Frontier definition

The primary benchmark is the frontier EFR defined as the 10th percentile of country EFRs within each GBD super-region × cause × year stratum (seven super-regions containing 5–46 countries each). Benchmarking within super-regions requires no external join to income or Socio-demographic Index data, avoids the circularity of adjusting a performance measure by a development covariate that is itself partly a function of health-system output, and respects broad regional epidemiological contexts such as pathogen mix, seasonality and malaria co-endemicity. The 10th percentile, rather than the minimum, is used so that no single country’s estimate (potentially distorted by small numbers or model artefacts at the favourable extreme) defines best practice for an entire region. This rationale holds only partially in the smallest stratum: South Asia contains just five countries, and percentile interpolation places its 10th-percentile frontier at or near the best-performing (minimum) country EFR, so the frontier there approximates a best-country definition rather than a best-decile one (see Limitations). The frontier is recomputed for every cause and year, so benchmarks evolve with observed performance rather than being fixed by assumption. Robustness was assessed with 5th- and 25th-percentile frontiers and with an aspirational global 10th-percentile frontier without regional stratification, the latter approximating what universal achievement of current global best practice would imply.

### Avoidable deaths

Avoidable deaths for country i are defined as avoidable_i = max(0, deaths_i − episodes_i × frontier_EFR_sr(i)), where frontier_EFR_sr(i) is the frontier for the super-region containing country i. The formula compares each country’s observed deaths with the deaths its own episode count would have generated at frontier performance; the zero floor ensures that countries at or below their frontier contribute zero rather than a negative credit. Estimates are summed by cause, super-region and year, and the avoidable share of deaths is reported alongside absolute counts so that composition effects can be distinguished from level effects. The frontier is relative, not normative: it measures the gap to the best decile currently achieved within a region, so estimates are conservative where an entire region lags global best practice.

### LRI incidence

Country-level LRI incidence for 1990 and 2019 was taken directly from GBD 2023 (ages 0–19 summed across the four age groups), and the 2023 cross-section was already fully observed, so all three anchor-year estimates rely on observed inputs only. For the full annual series shown in the trend figure, country episodes between the observed 1990, 2019 and 2023 anchors were linearly interpolated; the interpolation affects only the displayed trend line, not any anchor-year estimate.

### Uncertainty

Primary estimates are deterministic, computed from GBD central values and consistent with published GBD sums. Uncertainty intervals derive from 2,000 Monte Carlo draws, sampling deaths and episodes independently from lognormal distributions fitted to GBD 95% UIs while holding the frontier fixed at the point benchmark; countries are treated as independent, so intervals characterize input uncertainty rather than spatial covariance. For wide-UI causes (pertussis, URI), Monte Carlo medians exceed deterministic sums because of right-skewed draws interacting with the zero floor on avoidable deaths; we therefore report deterministic values as headline estimates, with Monte Carlo intervals reflecting input uncertainty, and flag wherever a point estimate falls outside its own interval. Lower and upper bounds for 0–19 sums are direct sums of age-group bounds (a workspace convention), not strict joint intervals.

### Population mapping (supplementary)

GLOBOCAN 2022 population estimates^12^ were mapped to 179 of 204 GBD countries (166 direct name matches plus 13 harmonized names); the 25 unmapped GBD countries—mostly small island states, Taiwan and Seychelles—account for 0.07% of the world 0–19 population. Population data were used only for supplementary denominators and played no role in EFR or avoidable-death computation.

### Sensitivity analyses

Six designs probe robustness: (a) frontier percentile (5th and 25th versus 10th); (b) an aspirational global 10th-percentile frontier providing an upper bound; (c) a pertussis frontier-pace counterfactual in which every country improves its EFR between 1990 and 2023 at the pace of its own super-region frontier, decomposing the residual gap into frontier stagnation versus country lag; (d) a 1990-frontier counterfactual applying each super-region’s 1990 frontier EFR to 2023 episodes to test whether 1990 best practice has been achieved globally; (e) an under-five age-band restriction in which the entire pipeline— country EFRs, super-region frontiers and avoidable deaths—is recomputed on the single GBD under-5-years age group (deaths and incident episodes), removing cross-country differences in 0–19 age structure by construction; and (f) development-gradient inequality: slope and concentration indices (SII/CII) of the country EFR and of the excess (avoidable) EFR, with countries ranked by GBD 2023 SDI (year-matched), ridits weighted by incident episodes of the corresponding year, and 95% confidence intervals from 1,000 bootstrap resamples over countries. In design (f), each country’s ridit is the cumulative episode-weighted share of the pooled country population below its SDI rank plus half of its own share; the SII is the episode-weighted least-squares slope of the outcome on this ridit, interpretable as the absolute difference in the outcome between the notional highest- and lowest-SDI ends of the ridit range; and the CII is the relative concentration index Σ p_i·y_i·(2r_i − 1)/μ, where p_i is country i’s episode-weight share, r_i its ridit, y_i its outcome and μ the weighted mean outcome. A paired bootstrap (5,000 resamples, seed 42, identical country indices drawn for both years) tests the 1990-to-2023 change in each index, and an equal-country-weight variant (1,000 resamples, seed 42) probes sensitivity to the episode weights.

## Results

### Global episode-fatality ratios and their dispersion

Table 1 summarizes deaths, episodes and EFRs by cause and anchor year. In 2023 the global 0–19 EFR was 8.78 deaths per 1,000 episodes for LRI (711,228 deaths among 81.0 million episodes), 6.11 per 1,000 for pertussis (112,839 deaths among 18.5 million episodes) and 2.9 per million for URI (16,377 deaths among 5.61 billion episodes). Between 1990 and 2023 the global LRI EFR fell 28% (12.26 to 8.78 per 1,000), while LRI deaths fell 65% because episodes roughly halved; the pertussis EFR fell from 9.19 to 6.11 per 1,000 over the same period. The URI EFR, three orders of magnitude lower, provides the near-zero-fatality contrast.

Country EFRs are widely dispersed within every super-region (Figure 1). In 2023 the LRI frontier EFR— the 10th percentile within each super-region—ranged from 0.73 per 1,000 episodes in the High-income super-region to 6.93 per 1,000 in Sub-Saharan Africa, a roughly tenfold spread in what the best decile of countries achieves even after regional stratification. Frontier trajectories also diverged: between 1990 and 2023 the High-income LRI frontier improved by about two-thirds (2.32 to 0.73 per 1,000), whereas the Sub-Saharan Africa frontier improved by only about 30% (9.85 to 6.93 per 1,000), and its pertussis frontier improved only 8.5% over 33 years.

**Table 1.** Global deaths, incident episodes and episode-fatality ratios (EFR) by cause, ages 0–19 years, 1990–2023.

| Cause | Year | Deaths (95% UI) | Episodes (95% UI) | EFR per 1,000 episodes |
|---|---|---|---|---|
| Lower respiratory infections | 1990 | 2,035,714 (1,556,792–2,615,533) | 166,066,423 (136,972,489–198,934,607) | 12.26 |
| Lower respiratory infections | 2019 | 816,491 (609,750–1,067,972) | 71,525,886 (59,280,541–85,257,366) | 11.42 |
| Lower respiratory infections | 2023 | 711,228 (507,283–958,451) | 80,998,085 (71,553,288–90,442,883) | 8.78 |
| Upper respiratory infections | 1990 | 27,128 (3,114–71,753) | 5,108,264,596 (3,792,251,736–6,683,247,660) | 0.00531 |
| Upper respiratory infections | 2019 | 17,290 (1,492–63,706) | 5,560,945,204 (4,103,834,698–7,299,182,756) | 0.00311 |
| Upper respiratory infections | 2023 | 16,377 (1,461–60,597) | 5,611,244,034 (4,107,753,095–7,411,233,413) | 0.00292 |
| Whooping cough (pertussis) | 1990 | 277,403 (31,138–753,341) | 30,194,376 (23,543,787–38,755,464) | 9.19 |
| Whooping cough (pertussis) | 2023 | 112,839 (12,545–321,874) | 18,469,250 (14,456,437–23,596,417) | 6.11 |

UI, uncertainty interval. EFR = deaths/incident episodes, expressed per 1,000 episodes (URI values fall below 0.01 per 1,000); note that EFR is not a clinical case-fatality ratio (see Methods). LRI episodes for 1990 and 2019 are observed GBD 2023 country-level estimates. Pertussis estimates unavailable for 2019.

**Figure 1.**
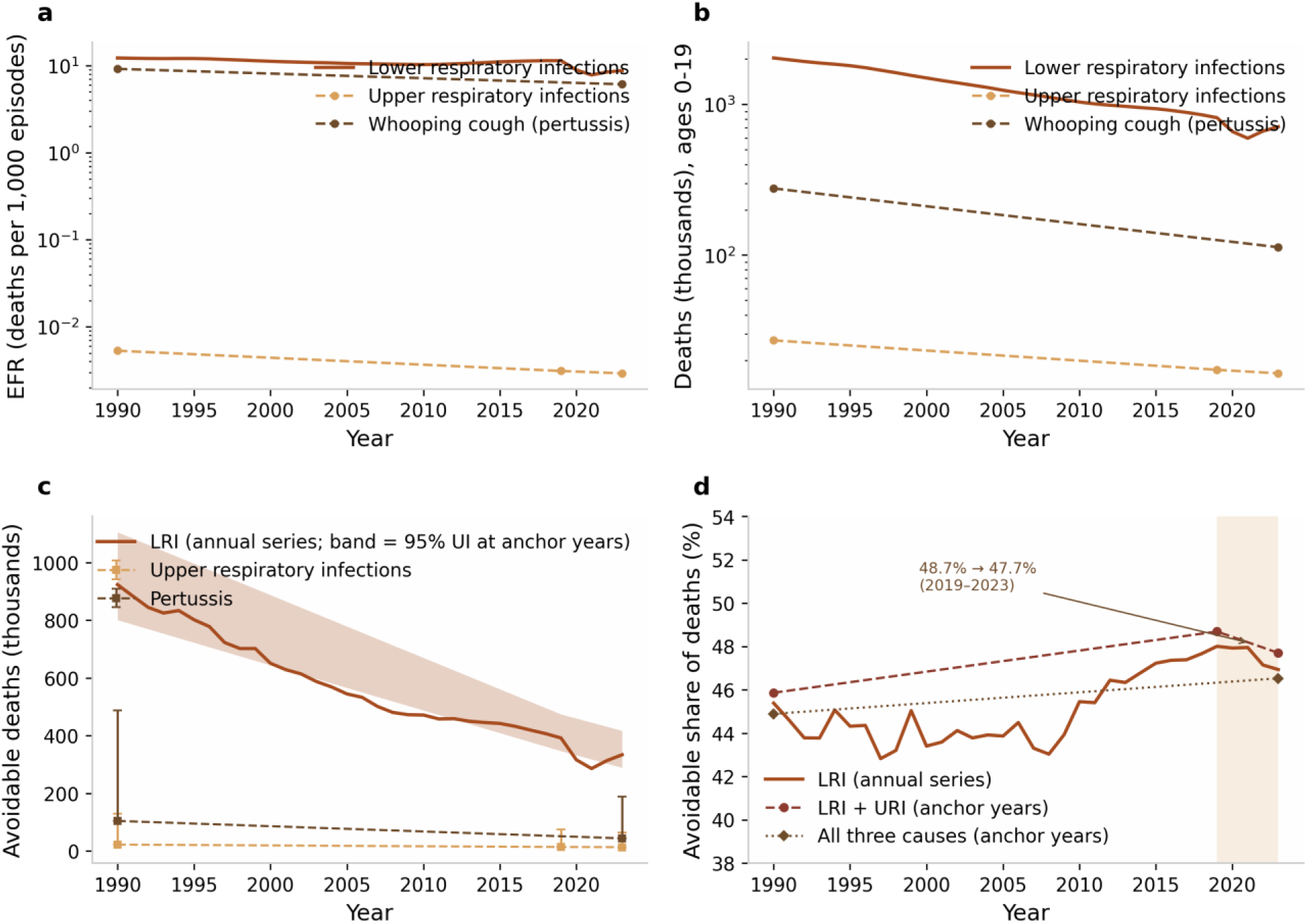
Global burden trends, ages 0–19 years. (a) Episode-fatality ratio (EFR, deaths per 1,000 episodes), 1990–2023: lower respiratory infections (LRI) as the full annual series; upper respiratory infections (URI) and pertussis at anchor years (1990, 2019, 2023; pertussis 1990 and 2023 only). (b) Deaths (thousands) by cause on the same temporal convention. (c) Avoidable deaths (thousands): LRI annual series with 95% Monte Carlo uncertainty band at anchor years; URI and pertussis deterministic estimates with Monte Carlo uncertainty intervals at anchor years. (d) Avoidable share of deaths (%): LRI annual series; LRI + URI and all three causes at anchor years. The shaded 2019–2023 pandemic window marks stalled convergence toward the frontier, with the avoidable share for LRI + URI edging from 48.7% to 47.7%.

### Avoidable deaths in 2023 and over time

Applying the super-region-stratified 10th-percentile frontier, 333,803 deaths from LRI (95% Monte Carlo UI 289,123–417,460; 46.9% of the 711,228 LRI deaths in 2023) were avoidable (Table 2). Adding the two secondary causes gives a deterministic combined total of 391,034 avoidable deaths (46.5% of the 840,444 deaths from the three causes); the combined figure is a deterministic sum, and a 95% UI is available for the LRI component only. The secondary-cause estimates are 43,958 (UI 49,473–187,891; 39.0% of pertussis deaths) for pertussis and 13,273 (UI 12,556–64,005; 81.0%) for URI. Both deterministic point estimates sit below their own Monte Carlo intervals, reflecting right-skewed input uncertainty interacting with the zero floor (see Methods), and the underlying GBD death estimates span a roughly 25-fold range for pertussis (12,545–321,874); these two figures should therefore be read as secondary results carrying wide uncertainty rather than as headline estimates.

The avoidable burden has contracted substantially in absolute terms but not as a share. In 1990, 1,050,468 deaths (44.9% of 2.34 million) were avoidable: LRI 923,895 (UI 801,022–1,105,630; 45.4%), pertussis 104,378 (37.6%) and URI 22,196 (81.8%) (rounded cause components sum to 1,050,469; the headline total is the rounded deterministic sum of unrounded values). By 2019, avoidable deaths for LRI+URI stood at 405,950 (48.7%; LRI 391,968, 48.0%; URI 13,982, 80.9%; pertussis estimates unavailable for 2019). Between 2019 and 2023, avoidable LRI+URI deaths fell 14.5% (405,950 to 347,076), tracking the overall mortality decline, and the avoidable *share* barely moved, edging downward (48.7% to 47.7%): mortality fell, yet the pattern is consistent with stalled convergence toward the frontier (Figure 2).

**Figure 2.**
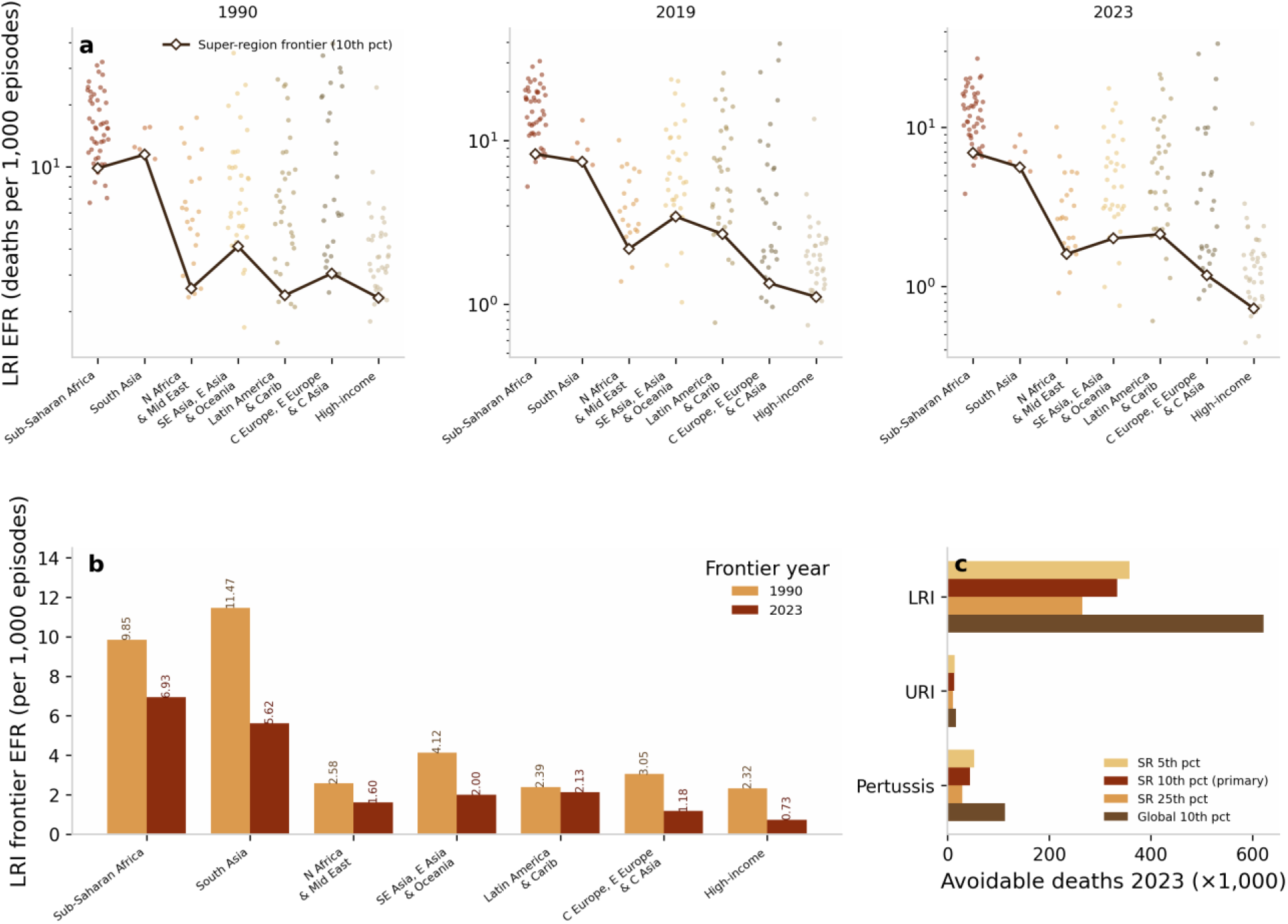
Frontier analysis of LRI episode-fatality ratios, ages 0–19 years. (a) Country EFRs (deaths per 1,000 episodes, log scale) by GBD super-region with the super-region 10th-percentile frontier, faceted by anchor year (1990, 2019, 2023). (b) LRI frontier EFR by super-region, 1990 versus 2023. (c) Sensitivity of 2023 avoidable-death estimates to the frontier definition: super-region 5th-, 10th-(primary) and 25th-percentile frontiers and the aspirational global 10th-percentile frontier, by cause.

### Geographic concentration and the poverty lock

The geography of avoidable death shifted markedly (Table 2; Figure 2A). In 1990 the burden was broadly distributed, with Southeast Asia, East Asia and Oceania holding the largest share; by 2023 Sub-Saharan Africa alone held 58.0% of all three-cause avoidable deaths, and Sub-Saharan Africa plus South Asia together held 73.1%, up from 41.8% in 1990. We refer to this pattern—the increasing concentration of the residual avoidable burden in low-income settings as other regions converge toward their frontiers—as a poverty lock. For URI, the near-zero-fatality contrast, Sub-Saharan Africa held 93.5% of avoidable deaths in 2023.

Concentration at the country level is sharper still (Table 3). The ten countries with the largest avoidable toll (Figure 4) account for 59.1% of 2023 avoidable deaths, and the top twenty for 77.1%. The five largest contributors are Nigeria (19.5%), India (13.8%), Niger (5.5%), the Democratic Republic of the Congo (4.5%) and Indonesia (3.3%). LRI dominates the avoidable total in every top-20 country, with pertussis contributing materially in Nigeria, Indonesia, Afghanistan, Angola and Somalia.

**Figure 3.**
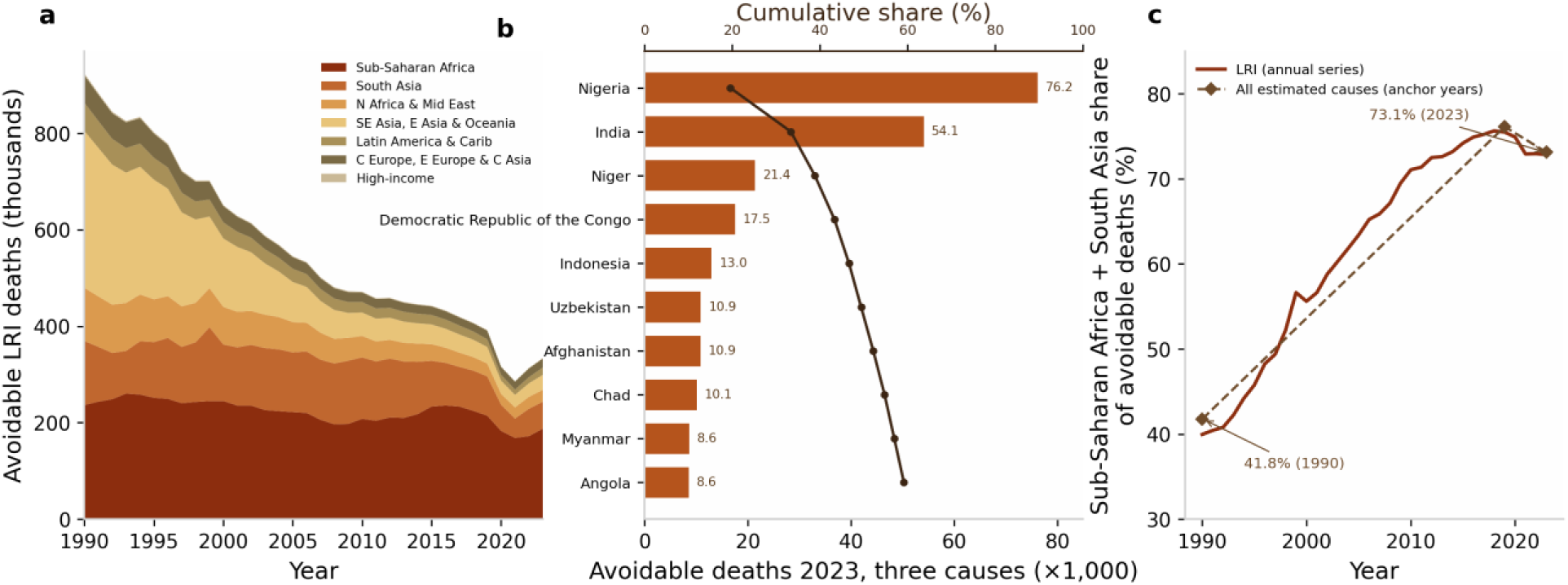
Geography of avoidable deaths, ages 0–19 years. (a) Avoidable LRI deaths by super-region, stacked annual series 1990–2023. (b) Top-10 countries by three-cause avoidable deaths in 2023, with the cumulative share of the global total (top 10 = 59.1%). (c) Combined Sub-Saharan Africa plus South Asia share of avoidable deaths: LRI annual series and all-estimated-cause anchor points (41.8% in 1990; 73.1% in 2023).

**Figure 4.**
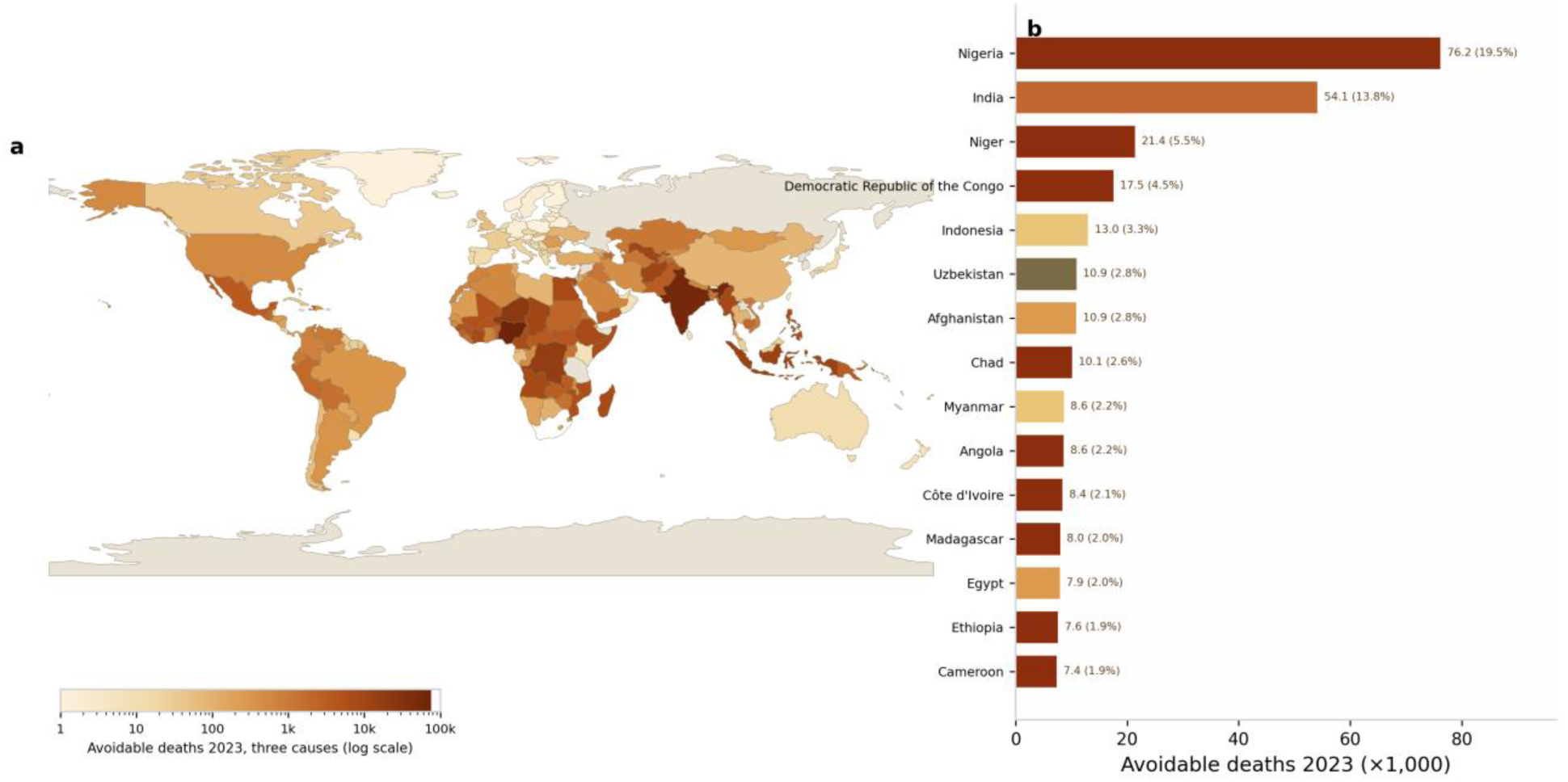
Country-level avoidable deaths in 2023, three causes combined, ages 0–19 years. (a) Choropleth map (logarithmic colour scale) on a Natural Earth 110 m basemap; 169 of 204 countries are represented and 35 small island states and territories not depicted on the basemap jointly account for 0.04% of avoidable deaths. (b) Top-15 countries ranked by avoidable deaths, with each country’s share of the global total in parentheses; bars are coloured by super-region.

### SDI-graded inequality of the fatality ratio

Ranking all 204 countries by SDI and weighting by incident episodes, the slope index of inequality for the LRI EFR was −12.5 deaths per 1,000 episodes (95% bootstrap CI −17.4 to −6.5) in 1990 and −12.9 (−18.3 to −6.8) in 2023, and the concentration index was −0.17 (−0.26 to −0.08) in 1990 and −0.24 (−0.39 to −0.12) in 2023: the fatality gradient across the development spectrum was steep in both years. For the excess (avoidable) EFR, the gradient was statistically indeterminate in 1990 (SII −2.8 per 1,000 episodes, −8.6 to 2.2; CII −0.08, −0.26 to 0.07) but significant in 2023 (SII −6.9 per 1,000, −11.0 to −2.6; CII −0.28, −0.45 to −0.11). A paired bootstrap test of the 1990-to-2023 change, however, does not confirm a significant shift for either outcome (excess EFR: ΔSII −4.1, −8.9 to 1.5, p = 0.175; ΔCII −0.195, −0.409 to 0.017, p = 0.083; LRI EFR: ΔSII p = 0.850; ΔCII p = 0.275): the two years’ intervals overlap and we do not claim a significant change, although the 2023 cross-section shows the avoidable component of case fatality distinctly concentrated in lower-SDI settings, consistent with the poverty-lock pattern (Table 5). The episode weights are heavy-tailed (Kish effective sample sizes 10.5 in 1990 and 14.6 in 2023; the five largest countries carry 56.3% and 49.0% of the total weight), so we repeated the estimation with equal country weights: the 1990 excess-EFR gradient is significant under equal weights (SII −9.6, −12.2 to −6.5; CII −0.27, −0.35 to −0.19), indicating that the 1990 indeterminacy is partly a property of the weighting. The excess-EFR indices are also truncated at zero by construction (24 countries in each year, at the high-SDI end), so they should not be compared directly with the all-in EFR indices (Table 5 note).

**Table 2.**
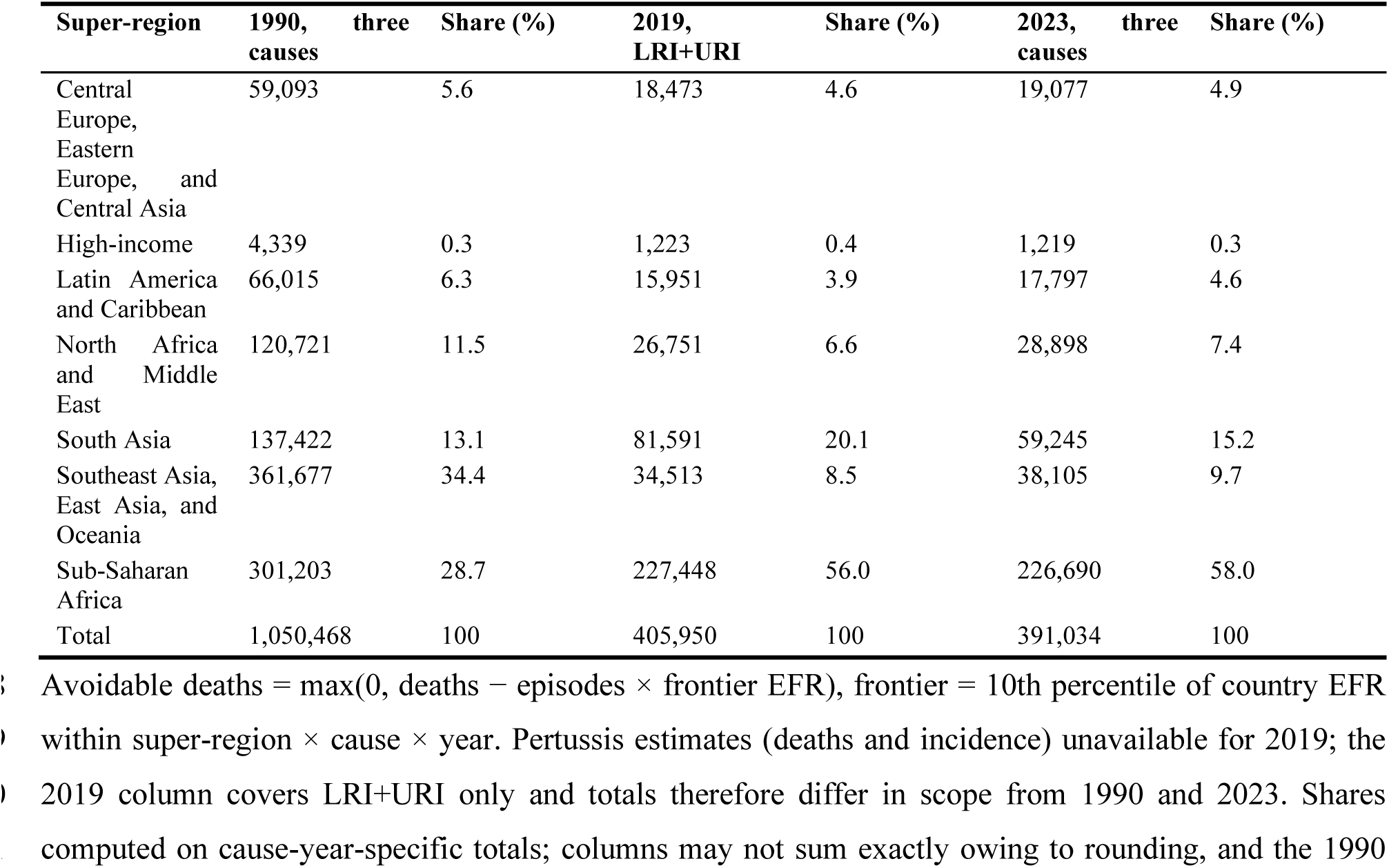
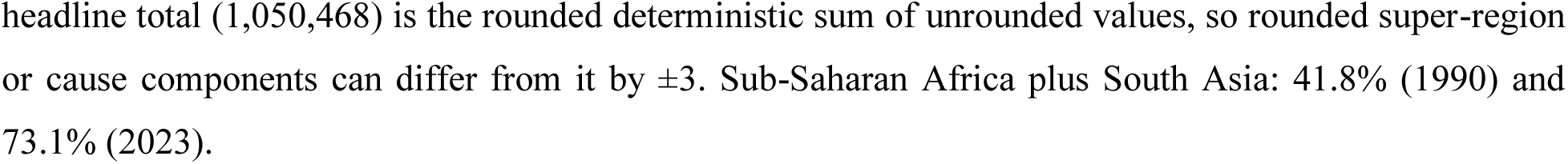
Avoidable deaths by super-region and year, ages 0–19 years.

| **Super-region** | **1990, three causes** | **Share (%)** | **2019, LRI+URI** | **Share (%)** | **2023, three causes** | **Share (%)** |
|---|---|---|---|---|---|---|
| Central Europe, Eastern Europe, and Central Asia | 59,093 | 5.6 | 18,473 | 4.6 | 19,077 | 4.9 |
| High-income | 4,339 | 0.3 | 1,223 | 0.4 | 1,219 | 0.3 |
| Latin America and Caribbean | 66,015 | 6.3 | 15,951 | 3.9 | 17,797 | 4.6 |
| North Africa and Middle East | 120,721 | 11.5 | 26,751 | 6.6 | 28,898 | 7.4 |
| South Asia | 137,422 | 13.1 | 81,591 | 20.1 | 59,245 | 15.2 |
| Southeast Asia, East Asia, and Oceania | 361,677 | 34.4 | 34,513 | 8.5 | 38,105 | 9.7 |
| Sub-Saharan Africa | 301,203 | 28.7 | 227,448 | 56.0 | 226,690 | 58.0 |
| Total | 1,050,468 | 100 | 405,950 | 100 | 391,034 | 100 |

Avoidable deaths = max(0, deaths − episodes × frontier EFR), frontier = 10th percentile of country EFR within super-region × cause × year. Pertussis estimates (deaths and incidence) unavailable for 2019; the 2019 column covers LRI+URI only and totals therefore differ in scope from 1990 and 2023. Shares computed on cause-year-specific totals; columns may not sum exactly owing to rounding, and the 1990 headline total (1,050,468) is the rounded deterministic sum of unrounded values, so rounded super-region or cause components can differ from it by ±3. Sub-Saharan Africa plus South Asia: 41.8% (1990) and 73.1% (2023).

**Table 3.**
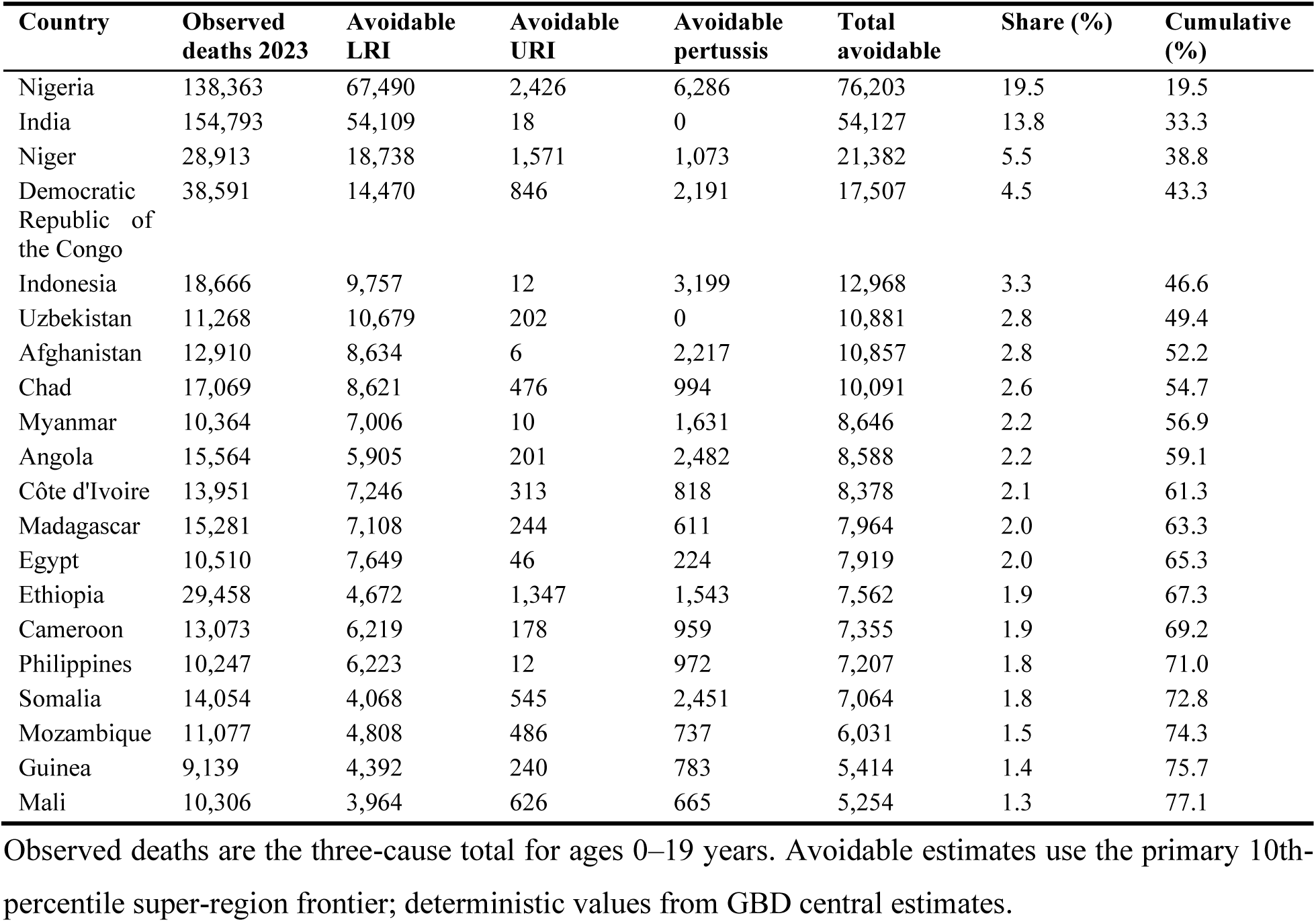
Top-20 countries by total avoidable deaths, three causes combined, 2023.

### Sensitivity analyses

Avoidable-death estimates are moderately sensitive to the frontier percentile but qualitatively stable (Table 4; Figure 3). For LRI in 2023, avoidable deaths range from 264,744 under the 25th-percentile frontier to 358,246 under the 5th-percentile frontier, bracketing the primary estimate of 333,803; corresponding ranges are 29,360–52,385 for pertussis and 10,214–14,136 for URI. The aspirational global 10th-percentile frontier, with no regional stratification, yields 621,483 avoidable LRI deaths (87% of LRI deaths) and 112,839 avoidable pertussis deaths (essentially 100%, because several high-coverage countries record near-zero pertussis fatality). The super-region-stratified frontier is therefore the conservative benchmark and the global frontier an upper bound. URI bounds the method at the near-zero-fatality extreme: with frontier EFRs near zero everywhere, about 81% of URI deaths are classified as avoidable.

The pertussis counterfactuals locate the residual gap. Had every country improved its EFR between 1990 and 2023 at the pace of its own super-region frontier, 2023 pertussis deaths would have been 140,055, *higher* than the observed 112,839, because the Sub-Saharan Africa frontier itself improved only 8.5% while most countries outperformed it. Countries lagging their regional frontier pace account for only 8,762 excess deaths (7.8% of observed). Conversely, had all countries achieved their super-region’s 1990 frontier EFR in 2023, deaths would have been 104,392, close to the observed value: 1990 best practice has been largely attained globally, and the remaining avoidable deaths reflect within-region lag behind 2023 best practice combined with stagnation of the frontier itself.

Restricting the analysis to the single under-five age band—the group in which 86% of the 0–19 LRI deaths of 2023 occur—removes cross-country differences in 0–19 age structure by construction and leaves every qualitative conclusion unchanged (Supplementary Table S7; Figure 5). The global under-5 LRI EFR in 2023 was 12.31 deaths per 1,000 episodes (8.78 for ages 0–19), and 297,328 under-5 LRI deaths (95% Monte Carlo UI 258,192–365,192) were avoidable, 48.8% of under-5 LRI deaths versus 46.9% for 0–19. The deterministic three-cause total was 344,500 avoidable deaths (47.9% of 718,676), versus 391,034 (46.5%) for 0–19, and the 1990 pattern was likewise preserved (1,023,856 avoidable deaths, 47.5%, versus 44.9%). Country rankings are essentially unaffected: the Spearman rank correlation between under-5 and 0–19 country avoidable-death totals in 2023 is 0.98 for the three causes combined and 0.95 for LRI alone (both P<0.001); the five largest contributors are identical and in the same order (Nigeria, India, Niger, the Democratic Republic of the Congo and Indonesia); and Sub-Saharan Africa plus South Asia hold 72.6% of under-5 avoidable deaths (73.1% for 0–19). Between 2019 and 2023, avoidable LRI+URI deaths in the under-5 band fell 19.1% while their share of deaths edged down from 52.2% to 49.6%, mirroring the 0–19 pattern (14.5% decline; 48.7% to 47.7%). Age-structure composition therefore explains neither the size of the avoidable burden nor its stalled convergence.

**Table 4.**
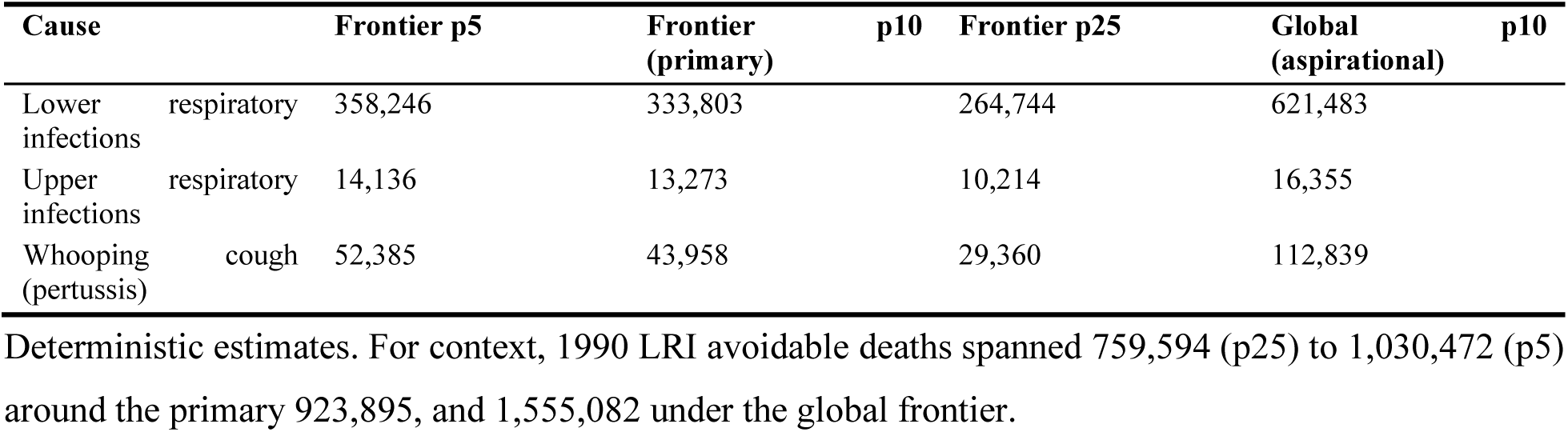
Sensitivity of avoidable-death estimates to frontier definition, 2023 (ages 0–19 years)

| **Cause** | **Frontier p5** | **Frontier p10 (primary)** | **Frontier p25** | **Global p10 (aspirational)** |
|---|---|---|---|---|
| Lower respiratory infections | 358,246 | 333,803 | 264,744 | 621,483 |
| Upper respiratory infections | 14,136 | 13,273 | 10,214 | 16,355 |
| Whooping cough (pertussis) | 52,385 | 43,958 | 29,360 | 112,839 |

Deterministic estimates. For context, 1990 LRI avoidable deaths spanned 759,594 (p25) to 1,030,472 (p5) around the primary 923,895, and 1,555,082 under the global frontier.

**Table 5.** Slope (SII) and concentration (CII) indices of inequality in the LRI episode-fatality ratio and the excess (avoidable) EFR, 204 countries ranked by SDI, 1990 and 2023.

| **Outcome (deaths per 1,000 episodes)** | **Year** | **SII (95% CI)** | **CII (95% CI)** |
|---|---|---|---|
| LRI EFR | 1990 | −12.5 (−17.4, −6.5) | −0.17 (−0.26, −0.08) |
| LRI EFR | 2023 | −12.9 (−18.3, −6.8) | −0.24 (−0.39, −0.12) |
| Excess (avoidable) EFR | 1990 | −2.8 (−8.6, 2.2) | −0.08 (−0.26, 0.07) |
| Excess (avoidable) EFR | 2023 | −6.9 (−11.0, −2.6) | −0.28 (−0.45, −0.11) |
| LRI EFR, change 2023 − 1990 (paired bootstrap) | 1990→2023 | Δ −0.4 (−8.3, 5.9), p = 0.850 | Δ −0.075 (−0.257, 0.048), p = 0.275 |
| Excess (avoidable) EFR, change 2023 − 1990 (paired bootstrap) | 1990→2023 | Δ −4.1 (−8.9, 1.5), p = 0.175 | Δ −0.195 (−0.409, 0.017), p = 0.083 |
| LRI EFR (equal country weights) | 1990 | −17.3 (−20.0, −14.0) | −0.276 (−0.319, −0.225) |
|  | 2023 | −14.6 (−16.6, −12.4) | −0.368 (−0.412, −0.317) |
| Excess (avoidable) EFR (equal country weights) | 1990 | −9.6 (−12.2, −6.5) | −0.274 (−0.352, −0.186) |
|  | 2023 | −8.5 (−10.2, −6.5) | −0.367 (−0.435, −0.290) |

Countries ranked by GBD 2023 SDI of the corresponding year; ridits weighted by incident LRI episodes of the same year (the at-risk denominator of the EFR, available for all 204 countries in both years). SII = episode-weighted least-squares slope of the outcome on the SDI ridit (absolute difference, deaths per 1,000 episodes, between the notional highest- and lowest-SDI ends of the ridit range); CII = concentration index (relative inequality; negative values indicate concentration in lower-SDI countries). Excess (avoidable) EFR = avoidable deaths/incident episodes, where avoidable deaths = max(0, deaths − episodes × frontier EFR). 95% CIs from 1,000 bootstrap resamples over countries (seed 42). Δ rows: paired bootstrap of the 1990-to-2023 change (5,000 resamples, seed 42, identical country indices drawn for both years); two-sided p = twice the smaller tail share of the Δ distribution. Equal-weight rows assign every country weight 1. The episode weights are heavy-tailed: Kish effective sample sizes are 10.5 (1990) and 14.6 (2023), and the five largest countries carry 56.3% (1990) and 49.0% (2023) of the total weight. The excess-EFR outcome is truncated at zero by construction (24 countries with zero avoidable deaths in each year; their mean SDI ridit is 0.82 in 1990 and 0.84 in 2023), which biases its indices toward zero, so the excess-EFR and all-in EFR indices should not be compared directly.

**Figure 5.**
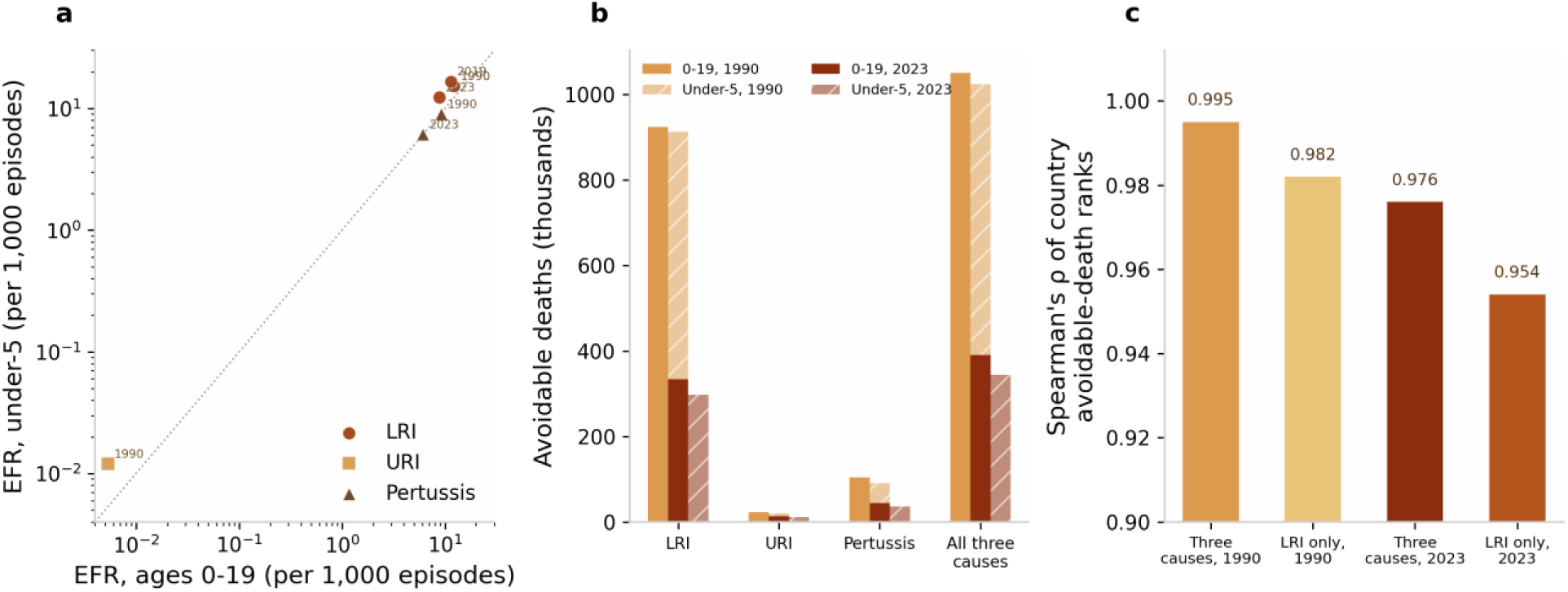
Under-5 versus 0–19 sensitivity analysis (identical EFR-frontier pipeline recomputed on under-5 estimates). (a) Under-5 EFR versus 0–19 EFR by cause and anchor year (log–log; dotted line, identity). (b) Avoidable deaths under-5 versus 0–19 by cause, 1990 and 2023. (c) Spearman rank concordance between under-5 and 0–19 country avoidable-death rankings (ρ = 0.954–0.995).

### Discussion Principal findings

Three decades after the World Summit for Children, nearly half of all deaths from childhood respiratory infections remain avoidable relative to episode-fatality ratios already achieved by the best-performing decile of countries within the same super-region: 333,803 LRI deaths in 2023 (95% UI 289,123–417,460), rising to a deterministic three-cause sum of 391,034 of 840,444 deaths for which a UI is available only for the LRI component. The absolute avoidable burden has fallen by nearly two-thirds since 1990, from 1,050,468 deaths, yet the avoidable share has not narrowed at all (44.9% in 1990 versus 46.5% in 2023), and during the 2019–2023 pandemic window it edged downward for LRI+URI (48.7% to 47.7%) even as absolute avoidable deaths fell 14.5%. This post-2019 mortality decline was carried by the fatality side, not the exposure side: LRI episodes rose by 13.2% between 2019 and 2023 while the LRI episode-fatality ratio fell by 23.1% (Table 1). Pandemic-era disruption and the asynchronous resurgence of respiratory virus circulation are part of the background to this exposure rebound,^13^ but over the same period the average country’s distance from its own regional frontier did not narrow: the pattern is consistent with stalled convergence.

This finding complicates a prevailing narrative of the Sustainable Development Goals era: that the “low-hanging fruit” of child survival has been harvested and that residual mortality is increasingly refractory, concentrated in neonatal conditions and complex systemic failures.^5^ For respiratory infections, the avoidable share of deaths in 2023 (46.5%) is no smaller than it was in 1990 (44.9%). The fruit has not been exhausted; rather, the tree has moved. Progress over three decades came overwhelmingly from fewer children falling ill—LRI episodes roughly halved—while the probability of dying once ill improved far more slowly, and the Sub-Saharan Africa LRI frontier improved by only about 30% in 33 years.

Two implications follow. First, exposure reduction and case survival should be monitored separately: aggregate mortality trends can mask a stagnant case-survival component when incidence is falling, and conversely can credit exposure reduction for gains that in fact came from better survival once ill. The EFR decomposition shows that the post-2019 decline belongs to the second pattern: episodes rose while fatality per episode fell. Second, the persistence of a near-constant avoidable share across three decades of expanding intervention coverage suggests that the marginal returns to coverage expansion alone are diminishing relative to the returns to the effectiveness of care delivered, a pattern the HAQ literature has documented at the level of composite indices^9^ and that our episode-based results now show within a single clinical pathway.

### Methodological contribution

The EFR frontier approach builds on the amenable-mortality tradition descending from Rutstein^6^ and Nolte and McKee,^7^ and on the mortality-to-incidence ratio literature in cancer epidemiology, where a deaths-to-cases ratio has long served as a system-level survival proxy and where its limitations have been examined in detail.^10^ Its increment over that lineage is specific and threefold: the denominator is episodes of illness rather than population, so avoidable deaths are estimated net of exposure and prevention and treatment contributions become separable; the benchmark is an empirical frontier of contemporaneous countries within the same super-region, requiring no external development covariate (the HAQ Index already uses an empirical frontier, but conditions it on development level^9^); and the metric is applied to respiratory infections, a cause group for which both deaths and incident episodes are modelled annually in GBD, allowing routine updating and extension to other such causes.^111^ The approach complements rather than replaces the HAQ Index and the Commission’s 8.6-million amenable-death accounting^89^: those frameworks map the total amenable envelope across dozens of causes, whereas the EFR frontier decomposes a single clinical pathway (episode onset to death) at country-year resolution. The URI contrast demonstrates both the method’s floor behaviour and its face validity: for a cause that is almost never fatal where care functions, about 81% of residual deaths are classified as avoidable, concentrated overwhelmingly in one super-region.

### Policy implications

The concentration of the avoidable burden is an actionable finding. Ten countries account for 59.1% of avoidable deaths, and Sub-Saharan Africa plus South Asia for 73.1%—up from 41.8% in 1990. For financing and delivery agencies, this means that a bounded set of national programmes, rather than a diffuse global effort, could in principle reach the majority of the avoidable burden with the established basic care package for childhood respiratory illness: timely access to effective antimicrobials, oxygen systems and pulse oximetry, supportive care for severe pneumonia, and catch-up immunization against pertussis and other respiratory pathogens where coverage has lapsed.^2,4,14^ None of these elements is technologically novel; the estimates presented here quantify the mortality cost of their incomplete and uneven deployment and provide a baseline against which scale-up can be monitored. The poverty lock is equally relevant for target-setting: because the residual burden is concentrated where fiscal space is narrowest, closing the frontier gap in high-burden countries will depend on sustained external financing for commodities—oxygen plants, antimicrobial supply chains and cold-chain capacity—whose returns are measurable in the very metric proposed here. That the avoidable *share* has plateaued even as coverage of several child-survival interventions expanded is consistent with the High Quality Health Systems Commission’s central argument—that poor-quality care, more than non-contact with services, now drives amenable mortality.^8^ Our estimates cannot observe quality directly, but an EFR that remains far above the regional frontier despite episode-level exposure declining is precisely the signature one would expect where children reach services yet receive sub-optimal treatment. We deliberately refrain from ranking or judging individual countries’ governance; the frontier is a benchmark for locating opportunity, not a scorecard of failure. The inequality indices describe the same pattern, although the two years’ intervals overlap and we do not claim a significant change: the excess-EFR concentration index was statistically indeterminate in 1990 and reached −0.28 (−0.45 to −0.11) in 2023, while the all-in EFR gradient across the SDI spectrum held at roughly thirteen deaths per 1,000 episodes in both years.

### Frontier stagnation versus country lag

The pertussis counterfactuals sharpen this diagnosis. Only 7.8% of observed 2023 pertussis deaths (8,762 of 112,839) are attributable to countries falling behind the pace of their own regional frontier; most countries matched or outperformed it. The residual avoidable burden exists because the frontier itself moved too slowly—Sub-Saharan Africa’s pertussis frontier improved just 8.5% in 33 years—while 1990 best practice (104,392 counterfactual deaths versus 112,839 observed) has effectively already been achieved. The policy implication is a shift in emphasis: catch-up interventions for laggards remain necessary but are no longer sufficient. Further large reductions require frontier-advancing investment— reliable oxygen supply chains, antimicrobial stewardship-compatible treatment access, neonatal and infant vaccination timeliness, and surveillance capable of detecting the pertussis resurgence now documented in the United States^15^—that raises the performance of the best decile itself, after which the convergence mechanism can propagate gains.

### Limitations

Six limitations qualify these results. First, the EFR is not a clinical case-fatality ratio: GBD episode denominators include mild community and outpatient illness (5.61 billion URI and 81.0 million LRI episodes in 2023), so EFRs are orders of magnitude below facility-based fatality rates and must not be compared with clinical CFRs.11 Second, all inputs are GBD 2023 modelled estimates, not vital-registration counts; for pertussis and URI the underlying death estimates carry very wide uncertainty intervals (global pertussis 2023: 12,545–321,874), which dominate the uncertainty of avoidable totals and explain why deterministic point estimates for these causes sit below their Monte Carlo intervals, and no uncertainty interval is available for the combined three-cause total. Third, the frontier definition is arbitrary in degree: the 10th percentile balances robustness against ambition, and estimates range from 264,744 to 358,246 avoidable LRI deaths across plausible percentile choices, with an aspirational global frontier bounding the total at 621,483. The frontier is also relative, so where an entire super-region lags, as the pertussis counterfactual shows for Sub-Saharan Africa, within-region benchmarking understates what global best practice would imply; and in South Asia, which contains only five countries, the 10th percentile falls close to the minimum country EFR, so the frontier there approximates a best-country definition and the 621,483 global-frontier estimate should be read as the relevant upper-bound contrast for that region. Fourth, the EFR is a crude ratio across the full 0–19 age band and is not age-standardized: because LRI fatality is concentrated in children under five, high-fertility countries whose 0–19 populations are structurally younger will record systematically higher EFRs than countries with older child populations even at identical age-specific fatality, inflating their estimated avoidable share. We addressed this concern directly with the under-five single-age-band sensitivity analysis, in which the composition effect is removed by construction: the avoidable share of deaths (47.9% versus 46.5% for the three causes in 2023), the country ranking (Spearman ρ 0.95–0.98) and the regional concentration (72.6% versus 73.1% in Sub-Saharan Africa plus South Asia) were all essentially unchanged, so age-structure confounding does not account for the headline findings. Residual composition effects within the under-five band (for example, the neonatal share of deaths) remain possible but operate over a much narrower age span. Fifth, the EFR reflects the severity mix of episodes as well as the quality of care: where malnutrition, HIV, comorbidity or incomplete vaccination raises the proportion of severe episodes, the fatality risk per episode is structurally higher, so the EFR is not a pure measure of case-management quality. Sixth, the analysis is ecological: country-level deaths and episodes cannot distinguish within-country variation by income, geography or facility quality, and avoidable-death counts should be interpreted as system-level opportunity estimates rather than individually preventable deaths. Supplementary population denominators were available for 179 of 204 countries (99.93% of the world 0– 19 population) and did not enter the core computation.

## Conclusions

Almost one in two deaths from childhood respiratory infections in 2023 (a deterministic three-cause sum of 391,034 deaths; 333,803 for LRI alone, 95% UI 289,123–417,460) would not have occurred had every country achieved the episode-fatality ratio of the best-performing decile within its own region. The absolute avoidable burden has fallen by nearly two-thirds since 1990, but the avoidable share has barely moved over three decades, the 2019–2023 pattern is consistent with stalled convergence toward the frontier, and the residual burden is now locked into Sub-Saharan Africa and South Asia (73.1% of avoidable deaths, up from 41.8% in 1990) and into ten countries holding 59.1%. In the pertussis counterfactual, most countries kept pace with their regional frontier, with only 7.8% of deaths attributable to laggards; the binding constraint is therefore no longer only catching up but advancing the frontier itself: higher-quality case management, dependable oxygen and antimicrobial access, and sustained and timely immunization. These estimates are bounded by the modelled nature of GBD inputs and the deliberately conservative within-region benchmark; under an aspirational global frontier the avoidable LRI toll alone would approach nine in ten deaths. Even under the conservative benchmark, however, the central conclusion stands: the avoidable fraction of a leading cause of child death has not narrowed in a generation. An episode-fatality frontier, computable annually from standard GBD outputs and requiring no external development covariates, offers a transparent, peer-benchmarked instrument for tracking whether that constraint is finally being relieved, and for distinguishing, year by year, between progress that comes from fewer children falling ill and progress that comes from fewer ill children dying.

## Presentation

This work has not been presented at any scientific meeting.

## Data availability

All inputs are publicly available from the Global Burden of Disease Study 2023 results tools and GLOBOCAN 2022; analytic tables accompanying this article list all country-year estimates. Version 3 additionally provides the inequality-analysis script (sii_cii_sdi_inequality.py), the country-year input frame (country_lri_efr_sdi_1990_2023.csv, including GBD location IDs) and its results (Supplementary_SII_CII_AvoidableRespDeaths.csv) in the accompanying code folder; SDI values are from the official covariate extraction IHME_GBD_2023_SDI_1950_2023_Y2025M10D12.CSV.

