## Supplementary_Materials for "Avoidable childhood respiratory-infection deaths: a frontier analysis of episode-fatality ratios in 204 countries, 1990–2023"

### Table S1. Population mapping rules: GLOBOCAN 2022 areas to GBD countries (237 GLOBOCAN labels)

| GLOBOCAN label | ISO3 | Population ages 0–19 | GBD country name | Mapping action |
|---|---|---|---|---|
| Afghanistan | AFG | 21,357,773 | Afghanistan | direct match |
| Albania | ALB | 667,552 | Albania | direct match |
| Algeria | DZA | 17,188,869 | Algeria | direct match |
| Angola | AGO | 19,908,256 | Angola | direct match |
| Azerbaijan | AZE | 3,048,614 | Azerbaijan | direct match |
| Argentina | ARG | 14,654,297 | Argentina | direct match |
| Australia | AUS | 6,608,989 | Australia | direct match |
| Austria | AUT | 1,759,419 | Austria | direct match |
| Bahamas | BHS | 116,135 | Bahamas | direct match |
| Bahrain | BHR | 420,926 | Bahrain | direct match |
| Bangladesh | BGD | 58,784,001 | Bangladesh | direct match |
| Armenia | ARM | 793,195 | Armenia | direct match |
| Barbados | BRB | 65,560 | Barbados | direct match |
| Belgium | BEL | 2,627,019 | Belgium | direct match |
| Bhutan | BTN | 258,309 | Bhutan | direct match |
| Bolivia (Plurinational State of) | BOL | 4,673,915 | Bolivia (Plurinational State of) | direct match |
| Bosnia Herzegovina | BIH | 619,883 | Bosnia and Herzegovina | name-mapped |
| Botswana | BWA | 1,031,252 | Botswana | direct match |
| Brazil | BRA | 59,150,434 | Brazil | direct match |
| Belize | BLZ | 156,490 | Belize | direct match |
| Solomon Islands | SLB | 359,285 | Solomon Islands | direct match |
| Brunei Darussalam | BRN | 129,106 | Brunei Darussalam | direct match |
| Bulgaria | BGR | 1,334,586 | Bulgaria | direct match |
| Myanmar | MMR | 18,661,754 | Myanmar | direct match |
| Burundi | BDI | 6,976,489 | Burundi | direct match |
| Belarus | BLR | 2,104,170 | Belarus | direct match |
| Cambodia | KHM | 6,722,377 | Cambodia | direct match |
| Cameroon | CMR | 14,602,598 | Cameroon | direct match |
| Canada | CAN | 8,051,474 | Canada | direct match |
| Cape Verde | CPV | 203,156 | Cabo Verde | name-mapped |
| Central African Republic | CAF | 2,756,528 | Central African Republic | direct match |
| Sri Lanka | LKA | 6,670,702 | Sri Lanka | direct match |
| Chad | TCD | 9,968,733 | Chad | direct match |
| Chile | CHL | 4,857,479 | Chile | direct match |
| China | CHN | 317,671,474 | China | direct match |
| Colombia | COL | 15,199,871 | Colombia | direct match |
| Comoros | COM | 442,577 | Comoros | direct match |
| Congo, Republic of | COG | 2,971,120 | Congo | name-mapped |

| GLOBOCAN label | ISO3 | Population ages 0–19 | GBD country name | Mapping action |
|---|---|---|---|---|
| Congo, Democratic Republic of | COD | 53,443,630 | Democratic Republic of the Congo | name-mapped |
| Costa Rica | CRI | 1,411,178 | Costa Rica | direct match |
| Croatia | HRV | 780,434 | Croatia | direct match |
| Cuba | CUB | 2,383,632 | Cuba | direct match |
| Cyprus | CYP | 273,313 | Cyprus | direct match |
| Czechia | CZE | 2,219,109 | Czechia | direct match |
| Benin | BEN | 6,664,887 | Benin | direct match |
| Denmark | DNK | 1,287,037 | Denmark | direct match |
| Dominican Republic | DOM | 3,927,886 | Dominican Republic | direct match |
| Ecuador | ECU | 6,430,772 | Ecuador | direct match |
| El Salvador | SLV | 2,270,559 | El Salvador | direct match |
| Equatorial Guinea | GNQ | 687,801 | Equatorial Guinea | direct match |
| Ethiopia | ETH | 60,657,079 | Ethiopia | direct match |
| Eritrea | ERI | 1,882,303 | Eritrea | direct match |
| Estonia | EST | 284,680 | Estonia | direct match |
| Fiji | FJI | 338,316 | Fiji | direct match |
| Finland | FIN | 1,161,908 | Finland | direct match |
| France (metropolitan) | FRA | 15,313,999 | France | name-mapped |
| French Guyana | GUF | 128,031 | French Guyana | dropped (aggregate or non-GBD territory) |
| French Polynesia | PYF | 83,125 | French Polynesia | dropped (aggregate or non-GBD territory) |
| Djibouti | DJI | 377,847 | Djibouti | direct match |
| Gabon | GAB | 1,075,689 | Gabon | direct match |
| Georgia | GEO | 1,030,469 | Georgia | direct match |
| The Republic of the Gambia | GMB | 1,388,876 | Gambia | name-mapped |
| Gaza Strip and West Bank | PSE | 2,549,404 | Palestine | name-mapped |
| Germany | DEU | 15,828,495 | Germany | direct match |
| Ghana | GHA | 15,123,083 | Ghana | direct match |
| Greece | GRC | 1,886,820 | Greece | direct match |
| France, Guadeloupe | GLP | 100,442 | France, Guadeloupe | dropped (aggregate or non-GBD territory) |
| Guam | GUM | 53,693 | Guam | direct match |
| Guatemala | GTM | 7,973,041 | Guatemala | direct match |
| Guinea | GIN | 7,449,761 | Guinea | direct match |
| Guyana | GUY | 287,669 | Guyana | direct match |
| Haiti | HTI | 4,861,232 | Haiti | direct match |
| Honduras | HND | 4,060,586 | Honduras | direct match |
| Hungary | HUN | 1,871,694 | Hungary | direct match |
| Iceland | ISL | 87,490 | Iceland | direct match |
| India | IND | 483,241,690 | India | direct match |
| Indonesia | IDN | 93,964,435 | Indonesia | direct match |
| Iran, Islamic Republic of | IRN | 27,054,540 | Iran (Islamic Republic of) | name-mapped |
| Iraq | IRQ | 19,895,338 | Iraq | direct match |
| Ireland | IRL | 1,342,693 | Ireland | direct match |
| Israel | ISR | 3,157,626 | Israel | direct match |
| Italy | ITA | 10,488,792 | Italy | direct match |

| GLOBOCAN label | ISO3 | Population ages 0–19 | GBD country name | Mapping action |
|---|---|---|---|---|
| Côte d'Ivoire | CIV | 14,433,977 | Côte d'Ivoire | direct match |
| Jamaica | JAM | 914,799 | Jamaica | direct match |
| Japan | JPN | 20,935,474 | Japan | direct match |
| Kazakhstan | KAZ | 6,869,700 | Kazakhstan | direct match |
| Jordan | JOR | 4,294,640 | Jordan | direct match |
| Kenya | KEN | 27,345,446 | Kenya | direct match |
| Korea, Democratic People's Republic of | PRK | 6,916,103 | Democratic People's Republic of Korea | name-mapped |
| Korea, Republic of | KOR | 8,534,281 | Republic of Korea | name-mapped |
| Kuwait | KWT | 1,177,872 | Kuwait | direct match |
| Kyrgyzstan | KGZ | 2,712,942 | Kyrgyzstan | direct match |
| Lao People's Democratic Republic | LAO | 3,055,467 | Lao People's Democratic Republic | direct match |
| Lebanon | LBN | 2,135,987 | Lebanon | direct match |
| Lesotho | LSO | 908,171 | Lesotho | direct match |
| Latvia | LVA | 401,474 | Latvia | direct match |
| Liberia | LBR | 2,685,302 | Liberia | direct match |
| Libya | LBY | 2,486,519 | Libya | direct match |
| Lithuania | LTU | 534,303 | Lithuania | direct match |
| Luxembourg | LUX | 135,137 | Luxembourg | direct match |
| Madagascar | MDG | 14,671,260 | Madagascar | direct match |
| Malawi | MWI | 10,808,525 | Malawi | direct match |
| Malaysia | MYS | 10,250,193 | Malaysia | direct match |
| Maldives | MDV | 136,005 | Maldives | direct match |
| Mali | MLI | 12,399,991 | Mali | direct match |
| Malta | MLT | 84,593 | Malta | direct match |
| France, Martinique | MTQ | 80,490 | France, Martinique | dropped (aggregate or non-GBD territory) |
| Mauritania | MRT | 2,422,068 | Mauritania | direct match |
| Mauritius | MUS | 294,945 | Mauritius | direct match |
| Mexico | MEX | 44,281,404 | Mexico | direct match |
| Mongolia | MNG | 1,291,627 | Mongolia | direct match |
| Republic of Moldova | MDA | 835,529 | Republic of Moldova | direct match |
| Montenegro | MNE | 151,077 | Montenegro | direct match |
| Morocco | MAR | 12,930,711 | Morocco | direct match |
| Mozambique | MOZ | 18,146,633 | Mozambique | direct match |
| Oman | OMN | 1,490,736 | Oman | direct match |
| Namibia | NAM | 1,215,669 | Namibia | direct match |
| Nepal | NPL | 11,349,820 | Nepal | direct match |
| The Netherlands | NLD | 3,651,730 | Netherlands | name-mapped |
| New Caledonia | NCL | 83,803 | New Caledonia | dropped (aggregate or non-GBD territory) |
| Vanuatu | VUT | 154,178 | Vanuatu | direct match |
| New Zealand | NZL | 1,247,156 | New Zealand | direct match |
| Nicaragua | NIC | 2,555,054 | Nicaragua | direct match |
| Niger | NER | 15,747,076 | Niger | direct match |
| Nigeria | NGA | 116,662,939 | Nigeria | direct match |
| Norway | NOR | 1,265,428 | Norway | direct match |
| Pakistan | PAK | 101,327,877 | Pakistan | direct match |

| GLOBOCAN label | ISO3 | Population ages 0–19 | GBD country name | Mapping action |
|---|---|---|---|---|
| Panama | PAN | 1,519,170 | Panama | direct match |
| Papua New Guinea | PNG | 4,160,329 | Papua New Guinea | direct match |
| Paraguay | PRY | 2,729,850 | Paraguay | direct match |
| Peru | PER | 10,762,155 | Peru | direct match |
| Philippines | PHL | 43,376,919 | Philippines | direct match |
| Poland | POL | 7,529,596 | Poland | direct match |
| Portugal | PRT | 1,803,322 | Portugal | direct match |
| Guinea-Bissau | GNB | 1,073,923 | Guinea-Bissau | direct match |
| Timor-Leste | TLS | 651,320 | Timor-Leste | direct match |
| Puerto Rico | PRI | 602,097 | Puerto Rico | direct match |
| Qatar | QAT | 534,637 | Qatar | direct match |
| France, La Réunion | REU | 267,479 | France, La Réunion | dropped (aggregate or non-GBD territory) |
| Romania | ROU | 3,945,769 | Romania | direct match |
| Russian Federation | RUS | 34,601,194 | Russian Federation | direct match |
| Rwanda | RWA | 6,717,826 | Rwanda | direct match |
| Saint Lucia | LCA | 44,654 | Saint Lucia | direct match |
| Sao Tome and Principe | STP | 119,363 | Sao Tome and Principe | direct match |
| Saudi Arabia | SAU | 11,103,537 | Saudi Arabia | direct match |
| Senegal | SEN | 9,293,621 | Senegal | direct match |
| Serbia | SRB | 1,806,829 | Serbia | direct match |
| Sierra Leone | SLE | 4,206,372 | Sierra Leone | direct match |
| Singapore | SGP | 989,410 | Singapore | direct match |
| Slovakia | SVK | 1,117,931 | Slovakia | direct match |
| Viet Nam | VNM | 29,471,164 | Viet Nam | direct match |
| Slovenia | SVN | 409,337 | Slovenia | direct match |
| Somalia | SOM | 9,626,715 | Somalia | direct match |
| South Africa | ZAF | 22,355,838 | South Africa | direct match |
| Zimbabwe | ZWE | 7,980,427 | Zimbabwe | direct match |
| Spain | ESP | 8,880,730 | Spain | direct match |
| South Sudan | SSD | 5,989,000 | South Sudan | direct match |
| Sudan | SDN | 22,991,347 | Sudan | direct match |
| Suriname | SUR | 206,920 | Suriname | direct match |
| Eswatini | SWZ | 564,000 | Eswatini | direct match |
| Sweden | SWE | 2,372,423 | Sweden | direct match |
| Switzerland | CHE | 1,748,530 | Switzerland | direct match |
| Syrian Arab Republic | SYR | 7,549,657 | Syrian Arab Republic | direct match |
| Tajikistan | TJK | 4,567,543 | Tajikistan | direct match |
| Thailand | THA | 15,495,742 | Thailand | direct match |
| Togo | TGO | 4,410,626 | Togo | direct match |
| Trinidad and Tobago | TTO | 367,173 | Trinidad and Tobago | direct match |
| United Arab Emirates | ARE | 1,968,271 | United Arab Emirates | direct match |
| Tunisia | TUN | 3,711,822 | Tunisia | direct match |
| Türkiye | TUR | 26,622,655 | Türkiye | direct match |
| Turkmenistan | TKM | 2,391,929 | Turkmenistan | direct match |
| Uganda | UGA | 27,418,282 | Uganda | direct match |
| Ukraine | UKR | 8,945,344 | Ukraine | direct match |
| North Macedonia | MKD | 452,782 | North Macedonia | direct match |
| Egypt | EGY | 44,843,899 | Egypt | direct match |

| GLOBOCAN label | ISO3 | Population ages 0–19 | GBD country name | Mapping action |
|---|---|---|---|---|
| United Kingdom | GBR | 15,834,550 | United Kingdom | direct match |
| Tanzania, United Republic of | TZA | 34,122,667 | United Republic of Tanzania | name-mapped |
| United States of America | USA | 82,081,756 | United States of America | direct match |
| Burkina Faso | BFA | 12,132,086 | Burkina Faso | direct match |
| Uruguay | URY | 943,003 | Uruguay | direct match |
| Uzbekistan | UZB | 12,457,525 | Uzbekistan | direct match |
| Venezuela | VEN | 10,205,788 | Venezuela (Bolivarian Republic of) | name-mapped |
| Samoa | WSM | 94,247 | Samoa | direct match |
| Yemen | YEM | 15,155,826 | Yemen | direct match |
| Zambia | ZMB | 10,651,812 | Zambia | direct match |
| World | — | 2,597,405,216 | World | dropped (aggregate or non-GBD territory) |
| Africa | — | 706,825,233 | Africa | dropped (aggregate or non-GBD territory) |
| Latin America and the Caribbean | — | 208,212,655 | Latin America and the Caribbean | dropped (aggregate or non-GBD territory) |
| Northern America | — | 90,161,343 | Northern America | dropped (aggregate or non-GBD territory) |
| Eastern Asia | — | 355,348,959 | Eastern Asia | dropped (aggregate or non-GBD territory) |
| Europe | — | 158,282,507 | Europe | dropped (aggregate or non-GBD territory) |
| Oceania | — | 13,423,573 | Oceania | dropped (aggregate or non-GBD territory) |
| Eastern Africa | — | 244,547,329 | Eastern Africa | dropped (aggregate or non-GBD territory) |
| Middle Africa | — | 105,533,718 | Middle Africa | dropped (aggregate or non-GBD territory) |
| Northern Africa | — | 104,370,345 | Northern Africa | dropped (aggregate or non-GBD territory) |
| Southern Africa | — | 26,074,930 | Southern Africa | dropped (aggregate or non-GBD territory) |
| Western Africa | — | 226,298,911 | Western Africa | dropped (aggregate or non-GBD territory) |
| Caribbean | — | 13,754,252 | Caribbean | dropped (aggregate or non-GBD territory) |
| Central America | — | 64,227,482 | Central America | dropped (aggregate or non-GBD territory) |
| South-Eastern Asia | — | 222,767,887 | South-Eastern Asia | dropped (aggregate or non-GBD territory) |
| South Central Asia | — | 739,180,356 | South Central Asia | dropped (aggregate or non-GBD territory) |
| Western Asia | — | 103,202,703 | Western Asia | dropped (aggregate or non-GBD territory) |
| Eastern Europe | — | 64,504,922 | Eastern Europe | dropped (aggregate or non-GBD territory) |
| Northern Europe | — | 24,637,366 | Northern Europe | dropped (aggregate or non-GBD territory) |
| Southern Europe | — | 28,060,445 | Southern Europe | dropped (aggregate or non-GBD territory) |

| GLOBOCAN label | ISO3 | Population ages 0–19 | GBD country name | Mapping action |
|---|---|---|---|---|
| Western Europe | — | 41,079,774 | Western Europe | dropped (aggregate or non-GBD territory) |
| Australia-New Zealand | — | 7,856,145 | Australia-New Zealand | dropped (aggregate or non-GBD territory) |
| Melanesia | — | 5,095,911 | Melanesia | dropped (aggregate or non-GBD territory) |
| South America | — | 130,230,921 | South America | dropped (aggregate or non-GBD territory) |
| Asia | — | 1,420,499,905 | Asia | dropped (aggregate or non-GBD territory) |
| European Union (27) | — | 89,326,343 | European Union (27) | dropped (aggregate or non-GBD territory) |
| Micronesia | — | 211,310 | Micronesia | dropped (aggregate or non-GBD territory) |
| Polynesia | — | 260,207 | Polynesia | dropped (aggregate or non-GBD territory) |
| Sub-Saharan Africa | — | 602,454,888 | Sub-Saharan Africa | dropped (aggregate or non-GBD territory) |
| Micronesia/Polynesia | — | 471,517 | Micronesia/Polynesia | dropped (aggregate or non-GBD territory) |
| Sub-Saharan Africa Hub | — | 602,283,704 | Sub-Saharan Africa Hub | dropped (aggregate or non-GBD territory) |
| Caribbean hub | — | 2,239,890 | Caribbean hub | dropped (aggregate or non-GBD territory) |
| Latin America Hub | — | 200,592,171 | Latin America Hub | dropped (aggregate or non-GBD territory) |
| South, East and South-Eastern Asia Hub | — | 1,288,297,563 | South, East and South-Eastern Asia Hub | dropped (aggregate or non-GBD territory) |
| Northern Africa, Central and Western Asia Hub | — | 236,082,196 | Northern Africa, Central and Western Asia Hub | dropped (aggregate or non-GBD territory) |
| Pacific Islands Hub | — | 5,160,048 | Pacific Islands Hub | dropped (aggregate or non-GBD territory) |
| Very HDI country | — | 379,770,450 | Very HDI country | dropped (aggregate or non-GBD territory) |
| High HDI country | — | 755,350,847 | High HDI country | dropped (aggregate or non-GBD territory) |
| Medium HDI country | — | 836,945,781 | Medium HDI country | dropped (aggregate or non-GBD territory) |
| Low HDI country | — | 624,281,203 | Low HDI country | dropped (aggregate or non-GBD territory) |
| High HDI country (but China) | — | 437,679,373 | High HDI country (but China) | dropped (aggregate or non-GBD territory) |
| Medium HDI country (but India) | — | 353,704,091 | Medium HDI country (but India) | dropped (aggregate or non-GBD territory) |
| High income | — | 258,585,305 | High income | dropped (aggregate or non-GBD territory) |
| Upper middle income | — | 658,519,073 | Upper middle income | dropped (aggregate or non-GBD territory) |
| Lower middle income | — | 1,303,933,951 | Lower middle income | dropped (aggregate or non-GBD territory) |
| Low income | — | 364,527,722 | Low income | dropped (aggregate or non-GBD territory) |

| GLOBOCAN label | ISO3 | Population ages 0–19 | GBD country name | Mapping action |
|---|---|---|---|---|
| WHO Africa region (AFRO) | — | 609,468,011 | WHO Africa region (AFRO) | dropped (aggregate or non-GBD territory) |
| WHO Americas region (PAHO) | — | 298,054,996 | WHO Americas region (PAHO) | dropped (aggregate or non-GBD territory) |
| WHO East Mediterranean region (EMRO) | — | 314,985,881 | WHO East Mediterranean region (EMRO) | dropped (aggregate or non-GBD territory) |
| WHO Europe region (EURO) | — | 222,098,899 | WHO Europe region (EURO) | dropped (aggregate or non-GBD territory) |
| WHO South-East Asia region (SEARO) | — | 696,129,881 | WHO South-East Asia region (SEARO) | dropped (aggregate or non-GBD territory) |
| WHO Western Pacific region (WPRO) | — | 455,610,613 | WHO Western Pacific region (WPRO) | dropped (aggregate or non-GBD territory) |

GLOBOCAN 2022 population estimates (single year 2022) mapped to GBD country names. 166 direct name matches plus 13 harmonized names cover 179 of 204 GBD countries; 25 unmapped GBD countries (mostly small island states, Taiwan and Seychelles) account for 0.07% of the world 0–19 population. 58 GLOBOCAN labels were dropped (regional, income and WHO aggregates and 6 non-GBD overseas territories). Population data were used only for supplementary denominators and played no role in EFR or avoidable-death computation.

#### Table S2. Complete frontier-sensitivity results: avoidable deaths under alternative frontier definitions, by cause and year, ages 0–19 years

| Cause | Year | Super-region p5 frontier | Super-region p10 frontier (primary) | Super-region p25 frontier | Global p10 frontier (aspirational) |
|---|---|---|---|---|---|
| Lower respiratory infections | 1990 | 1,030,472 | 923,895 | 759,594 | 1,555,082 |
| Lower respiratory infections | 2019 | 414,987 | 391,968 | 318,826 | 706,442 |
| Lower respiratory infections | 2023 | 358,189 | 333,803 | 264,723 | 621,511 |
| Upper respiratory infections | 1990 | 23,006 | 22,196 | 17,311 | 27,073 |
| Upper respiratory infections | 2019 | 14,942 | 13,982 | 10,125 | 17,268 |
| Upper respiratory infections | 2023 | 14,136 | 13,273 | 10,214 | 16,355 |
| Whooping cough (pertussis) | 1990 | 134,740 | 104,378 | 70,339 | 277,385 |
| Whooping cough (pertussis) | 2023 | 52,385 | 43,958 | 29,360 | 112,839 |

Deterministic estimates computed from GBD 2023 central values. The primary benchmark is the 10th percentile of country episode-fatality ratios within each super-region × cause × year stratum. Pertussis estimates unavailable for 2019.

**Table S3. Avoidable deaths by cause, super-region and year, ages 0–19 years (complete results)**

| Cause | Year | Super-region | Deaths | Avoidable deaths | Share of cause-year avoidable total (%) |
|---|---|---|---|---|---|
| Lower respiratory infections | 1990 | Central Europe, Eastern Europe, and Central Asia | 70,859 | 57,011 | 6.2 |
| Lower respiratory infections | 1990 | High-income | 8,068 | 3,573 | 0.4 |
| Lower respiratory infections | 1990 | Latin America and Caribbean | 96,493 | 58,619 | 6.3 |
| Lower respiratory infections | 1990 | North Africa and Middle East | 149,597 | 110,360 | 11.9 |
| Lower respiratory infections | 1990 | South Asia | 658,318 | 132,888 | 14.4 |
| Lower respiratory infections | 1990 | Southeast Asia, East Asia, and Oceania | 541,436 | 325,192 | 35.2 |
| Lower respiratory infections | 1990 | Sub-Saharan Africa | 510,942 | 236,253 | 25.6 |
| Lower respiratory infections | 2019 | Central Europe, Eastern Europe, and Central Asia | 20,142 | 18,068 | 4.6 |
| Lower respiratory infections | 2019 | High-income | 2,234 | 1,191 | 0.3 |
| Lower respiratory infections | 2019 | Latin America and Caribbean | 23,442 | 15,834 | 4.0 |
| Lower respiratory infections | 2019 | North Africa and Middle East | 41,890 | 26,675 | 6.8 |
| Lower respiratory infections | 2019 | South Asia | 243,124 | 81,445 | 20.8 |
| Lower respiratory infections | 2019 | Southeast Asia, East Asia, and Oceania | 62,594 | 34,321 | 8.8 |
| Lower respiratory infections | 2019 | Sub-Saharan Africa | 423,066 | 214,434 | 54.7 |
| Lower respiratory infections | 2023 | Central Europe, Eastern Europe, and Central Asia | 20,247 | 18,331 | 5.5 |
| Lower respiratory infections | 2023 | High-income | 1,933 | 1,170 | 0.4 |
| Lower respiratory infections | 2023 | Latin America and Caribbean | 22,297 | 15,347 | 4.6 |
| Lower respiratory infections | 2023 | North Africa and Middle East | 36,065 | 24,838 | 7.4 |
| Lower respiratory infections | 2023 | South Asia | 196,859 | 55,806 | 16.7 |
| Lower respiratory infections | 2023 | Southeast Asia, East Asia, and Oceania | 51,179 | 30,956 | 9.3 |

| Cause | Year | Super-region | Deaths | Avoidable deaths | Share of cause-year avoidable total (%) |
|---|---|---|---|---|---|
| Lower respiratory infections | 2023 | Sub-Saharan Africa | 382,648 | 187,354 | 56.1 |
| Upper respiratory infections | 1990 | Central Europe, Eastern Europe, and Central Asia | 1,073 | 973 | 4.4 |
| Upper respiratory infections | 1990 | High-income | 245 | 156 | 0.7 |
| Upper respiratory infections | 1990 | Latin America and Caribbean | 1,329 | 1,243 | 5.6 |
| Upper respiratory infections | 1990 | North Africa and Middle East | 138 | 135 | 0.6 |
| Upper respiratory infections | 1990 | South Asia | 2,243 | 311 | 1.4 |
| Upper respiratory infections | 1990 | Southeast Asia, East Asia, and Oceania | 6,547 | 6,545 | 29.5 |
| Upper respiratory infections | 1990 | Sub-Saharan Africa | 15,554 | 12,833 | 57.8 |
| Upper respiratory infections | 2019 | Central Europe, Eastern Europe, and Central Asia | 415 | 405 | 2.9 |
| Upper respiratory infections | 2019 | High-income | 41 | 32 | 0.2 |
| Upper respiratory infections | 2019 | Latin America and Caribbean | 144 | 117 | 0.8 |
| Upper respiratory infections | 2019 | North Africa and Middle East | 78 | 76 | 0.5 |
| Upper respiratory infections | 2019 | South Asia | 822 | 146 | 1.0 |
| Upper respiratory infections | 2019 | Southeast Asia, East Asia, and Oceania | 193 | 192 | 1.4 |
| Upper respiratory infections | 2019 | Sub-Saharan Africa | 15,597 | 13,014 | 93.1 |
| Upper respiratory infections | 2023 | Central Europe, Eastern Europe, and Central Asia | 398 | 391 | 2.9 |
| Upper respiratory infections | 2023 | High-income | 39 | 31 | 0.2 |
| Upper respiratory infections | 2023 | Latin America and Caribbean | 138 | 116 | 0.9 |
| Upper respiratory infections | 2023 | North Africa and Middle East | 65 | 63 | 0.5 |
| Upper respiratory infections | 2023 | South Asia | 683 | 124 | 0.9 |
| Upper respiratory infections | 2023 | Southeast Asia, East Asia, and Oceania | 134 | 134 | 1.0 |

| Cause | Year | Super-region | Deaths | Avoidable deaths | Share of cause-year avoidable total (%) |
|---|---|---|---|---|---|
| Upper respiratory infections | 2023 | Sub-Saharan Africa | 14,920 | 12,413 | 93.5 |
| Whooping cough (pertussis) | 1990 | Central Europe, Eastern Europe, and Central Asia | 1,109 | 1,109 | 1.1 |
| Whooping cough (pertussis) | 1990 | High-income | 610 | 610 | 0.6 |
| Whooping cough (pertussis) | 1990 | Latin America and Caribbean | 6,155 | 6,154 | 5.9 |
| Whooping cough (pertussis) | 1990 | North Africa and Middle East | 19,477 | 10,226 | 9.8 |
| Whooping cough (pertussis) | 1990 | South Asia | 106,004 | 4,223 | 4.0 |
| Whooping cough (pertussis) | 1990 | Southeast Asia, East Asia, and Oceania | 52,113 | 29,940 | 28.7 |
| Whooping cough (pertussis) | 1990 | Sub-Saharan Africa | 91,935 | 52,117 | 49.9 |
| Whooping cough (pertussis) | 2023 | Central Europe, Eastern Europe, and Central Asia | 355 | 355 | 0.8 |
| Whooping cough (pertussis) | 2023 | High-income | 18 | 18 | 0.0 |
| Whooping cough (pertussis) | 2023 | Latin America and Caribbean | 2,335 | 2,334 | 5.3 |
| Whooping cough (pertussis) | 2023 | North Africa and Middle East | 5,934 | 3,997 | 9.1 |
| Whooping cough (pertussis) | 2023 | South Asia | 19,153 | 3,315 | 7.5 |
| Whooping cough (pertussis) | 2023 | Southeast Asia, East Asia, and Oceania | 10,850 | 7,015 | 16.0 |
| Whooping cough (pertussis) | 2023 | Sub-Saharan Africa | 74,194 | 26,923 | 61.2 |

Avoidable deaths = max(0, deaths − episodes × frontier EFR), frontier = 10th percentile of country EFR within super-region × cause × year. Pertussis estimates unavailable for 2019. Values rounded to whole deaths; totals of rounded components may differ by ±1 from rounded totals.

#### Table S4. Top-20 countries by total avoidable deaths, three causes combined, 2023 (complete detail)

| Country | Observed deaths 2023 (three causes) | Avoidable LRI | Avoidable URI | Avoidable pertussis | Total avoidable | Share (%) | Cumulative (%) |
|---|---|---|---|---|---|---|---|
| Nigeria | 138,363 | 67,490 | 2,426 | 6,286 | 76,203 | 19.5 | 19.5 |
| India | 154,793 | 54,109 | 18 | 0 | 54,127 | 13.8 | 33.3 |
| Niger | 28,913 | 18,738 | 1,571 | 1,073 | 21,382 | 5.5 | 38.8 |

| Country | Observed deaths 2023 (three causes) | Avoidable LRI | Avoidable URI | Avoidable pertussis | Total avoidable | Share (%) | Cumulative (%) |
|---|---|---|---|---|---|---|---|
| Democratic Republic of the Congo | 38,591 | 14,470 | 846 | 2,191 | 17,507 | 4.5 | 43.3 |
| Indonesia | 18,666 | 9,757 | 12 | 3,199 | 12,968 | 3.3 | 46.6 |
| Uzbekistan | 11,268 | 10,679 | 202 | 0 | 10,881 | 2.8 | 49.4 |
| Afghanistan | 12,910 | 8,634 | 6 | 2,217 | 10,857 | 2.8 | 52.2 |
| Chad | 17,069 | 8,621 | 476 | 994 | 10,091 | 2.6 | 54.7 |
| Myanmar | 10,364 | 7,006 | 10 | 1,631 | 8,646 | 2.2 | 56.9 |
| Angola | 15,564 | 5,905 | 201 | 2,482 | 8,588 | 2.2 | 59.1 |
| Côte d'Ivoire | 13,951 | 7,246 | 313 | 818 | 8,378 | 2.1 | 61.3 |
| Madagascar | 15,281 | 7,108 | 244 | 611 | 7,964 | 2.0 | 63.3 |
| Egypt | 10,510 | 7,649 | 46 | 224 | 7,919 | 2.0 | 65.3 |
| Ethiopia | 29,458 | 4,672 | 1,347 | 1,543 | 7,562 | 1.9 | 67.3 |
| Cameroon | 13,073 | 6,219 | 178 | 959 | 7,355 | 1.9 | 69.2 |
| Philippines | 10,247 | 6,223 | 12 | 972 | 7,207 | 1.8 | 71.0 |
| Somalia | 14,054 | 4,068 | 545 | 2,451 | 7,064 | 1.8 | 72.8 |
| Mozambique | 11,077 | 4,808 | 486 | 737 | 6,031 | 1.5 | 74.3 |
| Guinea | 9,139 | 4,392 | 240 | 783 | 5,414 | 1.4 | 75.7 |
| Mali | 10,306 | 3,964 | 626 | 665 | 5,254 | 1.3 | 77.1 |

Ages 0–19 years. Avoidable estimates use the primary 10th-percentile super-region frontier; deterministic values from GBD 2023 central estimates. The top 10 countries account for 59.1% and the top 20 for 77.1% of 2023 avoidable deaths.

### Section S5. Monte Carlo uncertainty methods and the LRI incidence series

Monte Carlo uncertainty. Primary avoidable-death estimates are deterministic, computed from GBD 2023 central values and consistent with published GBD sums. Uncertainty intervals derive from 2,000 Monte Carlo draws per country, cause and year. Deaths and incident episodes are sampled independently from lognormal distributions fitted to the GBD 95% uncertainty intervals

for each input, while the frontier EFR is held fixed at the point benchmark (the 10th percentile of country central-value EFRs within each super-region × cause × year stratum). Countries are treated as independent draws, so the intervals characterize input uncertainty rather than spatial covariance. For wide-UI causes (pertussis and upper respiratory infections), Monte Carlo medians exceed deterministic sums because right-skewed draws interact with the zero floor on avoidable deaths (a draw in which deaths fall below episodes × frontier EFR contributes zero, never a negative value). Deterministic values are therefore reported as headline estimates, with Monte Carlo intervals reflecting input uncertainty, and every point estimate that falls outside its own interval is flagged in the text. Lower and upper bounds for 0–19-year sums are direct sums of age-group bounds (a workspace convention), not strict joint intervals. Table S5 reports the Monte Carlo medians and 95% intervals for global avoidable deaths.

### Table S5. Monte Carlo results for global avoidable deaths, by cause and year

| Cause | Year | MC median avoidable deaths | 95% UI lower | 95% UI upper |
|---|---|---|---|---|
| Lower respiratory infections | 1990 | 945,633 | 801,022 | 1,105,630 |

| Cause | Year | MC median avoidable deaths | 95% UI lower | 95% UI upper |
|---|---|---|---|---|
| Lower respiratory infections | 2019 | 402,786 | 346,747 | 474,286 |
| Lower respiratory infections | 2023 | 347,175 | 289,123 | 417,460 |
| Upper respiratory infections | 1990 | 43,183 | 21,778 | 129,704 |
| Upper respiratory infections | 2019 | 26,311 | 13,588 | 75,719 |
| Upper respiratory infections | 2023 | 24,967 | 12,556 | 64,005 |
| Whooping cough (pertussis) | 1990 | 218,273 | 119,704 | 487,143 |
| Whooping cough (pertussis) | 2023 | 92,526 | 49,473 | 187,891 |

2,000 Monte Carlo draws; lognormal sampling of deaths and episodes fitted to GBD 95% UIs with the frontier fixed. Headline estimates in the main text are deterministic sums of GBD central values, not these medians.

LRI incidence series. Country-level LRI incidence for 1990, 2019 and 2023 is taken directly from GBD 2023 (ages 0–19 years, summed across the four age groups <5, 5–9, 10–14 and 15–19 years); the global 0–19 sums are 166,066,423 episodes in 1990 and 71,525,886 in 2019, within 0.1% of the published GBD global series. All three anchor-year estimates therefore rely on observed inputs only. For the full annual series shown in the trend figure (Figure 2A), country episodes between the observed 1990, 2019 and 2023 anchors are linearly interpolated; the interpolation affects only the displayed trend line, not any anchor-year estimate.

### Section S6. GLOBOCAN 2022 population denominators (supplementary use only)

GLOBOCAN 2022 population estimates (237 areas, single year 2022) were mapped to 179 of 204 GBD countries: 166 direct name matches plus 13 harmonized names (full mapping rules in Table S1). The 25 unmapped GBD countries—mostly small island states, Taiwan and Seychelles —account for 0.07% of the world 0–19 population. 58 GLOBOCAN labels were dropped because they are regional, income or WHO aggregates or non-GBD overseas territories. Population data were used only for supplementary denominators and played no role in the EFR or avoidable-death computation. Table S6 lists the mapped 0–19 population for each of the 179 GBD countries.

### Table S6. Population ages 0–19 years for the 179 mapped GBD countries (GLOBOCAN 2022)

| GBD country | ISO3 | Population ages 0–19 (2022) |
|---|---|---|
| Afghanistan | AFG | 21,357,773 |
| Albania | ALB | 667,552 |
| Algeria | DZA | 17,188,869 |
| Angola | AGO | 19,908,256 |
| Azerbaijan | AZE | 3,048,614 |
| Argentina | ARG | 14,654,297 |
| Australia | AUS | 6,608,989 |
| Austria | AUT | 1,759,419 |
| Bahamas | BHS | 116,135 |
| Bahrain | BHR | 420,926 |
| Bangladesh | BGD | 58,784,001 |

| GBD country | ISO3 | Population ages 0–19 (2022) |
|---|---|---|
| Armenia | ARM | 793,195 |
| Barbados | BRB | 65,560 |
| Belgium | BEL | 2,627,019 |
| Bhutan | BTN | 258,309 |
| Bolivia (Plurinational State of) | BOL | 4,673,915 |
| Bosnia and Herzegovina | BIH | 619,883 |
| Botswana | BWA | 1,031,252 |
| Brazil | BRA | 59,150,434 |
| Belize | BLZ | 156,490 |
| Solomon Islands | SLB | 359,285 |
| Brunei Darussalam | BRN | 129,106 |
| Bulgaria | BGR | 1,334,586 |
| Myanmar | MMR | 18,661,754 |
| Burundi | BDI | 6,976,489 |
| Belarus | BLR | 2,104,170 |
| Cambodia | KHM | 6,722,377 |
| Cameroon | CMR | 14,602,598 |
| Canada | CAN | 8,051,474 |
| Cabo Verde | CPV | 203,156 |
| Central African Republic | CAF | 2,756,528 |
| Sri Lanka | LKA | 6,670,702 |
| Chad | TCD | 9,968,733 |
| Chile | CHL | 4,857,479 |
| China | CHN | 317,671,474 |
| Colombia | COL | 15,199,871 |
| Comoros | COM | 442,577 |
| Congo | COG | 2,971,120 |
| Democratic Republic of the Congo | COD | 53,443,630 |
| Costa Rica | CRI | 1,411,178 |
| Croatia | HRV | 780,434 |
| Cuba | CUB | 2,383,632 |
| Cyprus | CYP | 273,313 |
| Czechia | CZE | 2,219,109 |
| Benin | BEN | 6,664,887 |
| Denmark | DNK | 1,287,037 |
| Dominican Republic | DOM | 3,927,886 |
| Ecuador | ECU | 6,430,772 |
| El Salvador | SLV | 2,270,559 |
| Equatorial Guinea | GNQ | 687,801 |
| Ethiopia | ETH | 60,657,079 |
| Eritrea | ERI | 1,882,303 |
| Estonia | EST | 284,680 |
| Fiji | FJI | 338,316 |
| Finland | FIN | 1,161,908 |
| France | FRA | 15,313,999 |
| Djibouti | DJI | 377,847 |
| Gabon | GAB | 1,075,689 |
| Georgia | GEO | 1,030,469 |

| GBD country | ISO3 | Population ages 0–19 (2022) |
|---|---|---|
| Gambia | GMB | 1,388,876 |
| Palestine | PSE | 2,549,404 |
| Germany | DEU | 15,828,495 |
| Ghana | GHA | 15,123,083 |
| Greece | GRC | 1,886,820 |
| Guam | GUM | 53,693 |
| Guatemala | GTM | 7,973,041 |
| Guinea | GIN | 7,449,761 |
| Guyana | GUY | 287,669 |
| Haiti | HTI | 4,861,232 |
| Honduras | HND | 4,060,586 |
| Hungary | HUN | 1,871,694 |
| Iceland | ISL | 87,490 |
| India | IND | 483,241,690 |
| Indonesia | IDN | 93,964,435 |
| Iran (Islamic Republic of) | IRN | 27,054,540 |
| Iraq | IRQ | 19,895,338 |
| Ireland | IRL | 1,342,693 |
| Israel | ISR | 3,157,626 |
| Italy | ITA | 10,488,792 |
| Côte d'Ivoire | CIV | 14,433,977 |
| Jamaica | JAM | 914,799 |
| Japan | JPN | 20,935,474 |
| Kazakhstan | KAZ | 6,869,700 |
| Jordan | JOR | 4,294,640 |
| Kenya | KEN | 27,345,446 |
| Democratic People's Republic of Korea | PRK | 6,916,103 |
| Republic of Korea | KOR | 8,534,281 |
| Kuwait | KWT | 1,177,872 |
| Kyrgyzstan | KGZ | 2,712,942 |
| Lao People's Democratic Republic | LAO | 3,055,467 |
| Lebanon | LBN | 2,135,987 |
| Lesotho | LSO | 908,171 |
| Latvia | LVA | 401,474 |
| Liberia | LBR | 2,685,302 |
| Libya | LBY | 2,486,519 |
| Lithuania | LTU | 534,303 |
| Luxembourg | LUX | 135,137 |
| Madagascar | MDG | 14,671,260 |
| Malawi | MWI | 10,808,525 |
| Malaysia | MYS | 10,250,193 |
| Maldives | MDV | 136,005 |
| Mali | MLI | 12,399,991 |
| Malta | MLT | 84,593 |
| Mauritania | MRT | 2,422,068 |
| Mauritius | MUS | 294,945 |
| Mexico | MEX | 44,281,404 |
| Mongolia | MNG | 1,291,627 |

| GBD country | ISO3 | Population ages 0–19 (2022) |
|---|---|---|
| Republic of Moldova | MDA | 835,529 |
| Montenegro | MNE | 151,077 |
| Morocco | MAR | 12,930,711 |
| Mozambique | MOZ | 18,146,633 |
| Oman | OMN | 1,490,736 |
| Namibia | NAM | 1,215,669 |
| Nepal | NPL | 11,349,820 |
| Netherlands | NLD | 3,651,730 |
| Vanuatu | VUT | 154,178 |
| New Zealand | NZL | 1,247,156 |
| Nicaragua | NIC | 2,555,054 |
| Niger | NER | 15,747,076 |
| Nigeria | NGA | 116,662,939 |
| Norway | NOR | 1,265,428 |
| Pakistan | PAK | 101,327,877 |
| Panama | PAN | 1,519,170 |
| Papua New Guinea | PNG | 4,160,329 |
| Paraguay | PRY | 2,729,850 |
| Peru | PER | 10,762,155 |
| Philippines | PHL | 43,376,919 |
| Poland | POL | 7,529,596 |
| Portugal | PRT | 1,803,322 |
| Guinea-Bissau | GNB | 1,073,923 |
| Timor-Leste | TLS | 651,320 |
| Puerto Rico | PRI | 602,097 |
| Qatar | QAT | 534,637 |
| Romania | ROU | 3,945,769 |
| Russian Federation | RUS | 34,601,194 |
| Rwanda | RWA | 6,717,826 |
| Saint Lucia | LCA | 44,654 |
| Sao Tome and Principe | STP | 119,363 |
| Saudi Arabia | SAU | 11,103,537 |
| Senegal | SEN | 9,293,621 |
| Serbia | SRB | 1,806,829 |
| Sierra Leone | SLE | 4,206,372 |
| Singapore | SGP | 989,410 |
| Slovakia | SVK | 1,117,931 |
| Viet Nam | VNM | 29,471,164 |
| Slovenia | SVN | 409,337 |
| Somalia | SOM | 9,626,715 |
| South Africa | ZAF | 22,355,838 |
| Zimbabwe | ZWE | 7,980,427 |
| Spain | ESP | 8,880,730 |
| South Sudan | SSD | 5,989,000 |
| Sudan | SDN | 22,991,347 |
| Suriname | SUR | 206,920 |
| Eswatini | SWZ | 564,000 |
| Sweden | SWE | 2,372,423 |

| GBD country | ISO3 | Population ages 0–19 (2022) |
|---|---|---|
| Switzerland | CHE | 1,748,530 |
| Syrian Arab Republic | SYR | 7,549,657 |
| Tajikistan | TJK | 4,567,543 |
| Thailand | THA | 15,495,742 |
| Togo | TGO | 4,410,626 |
| Trinidad and Tobago | TTO | 367,173 |
| United Arab Emirates | ARE | 1,968,271 |
| Tunisia | TUN | 3,711,822 |
| Türkiye | TUR | 26,622,655 |
| Turkmenistan | TKM | 2,391,929 |
| Uganda | UGA | 27,418,282 |
| Ukraine | UKR | 8,945,344 |
| North Macedonia | MKD | 452,782 |
| Egypt | EGY | 44,843,899 |
| United Kingdom | GBR | 15,834,550 |
| United Republic of Tanzania | TZA | 34,122,667 |
| United States of America | USA | 82,081,756 |
| Burkina Faso | BFA | 12,132,086 |
| Uruguay | URY | 943,003 |
| Uzbekistan | UZB | 12,457,525 |
| Venezuela (Bolivarian Republic of) | VEN | 10,205,788 |
| Samoa | WSM | 94,247 |
| Yemen | YEM | 15,155,826 |
| Zambia | ZMB | 10,651,812 |

Source: GLOBOCAN 2022 (Bray et al., CA Cancer J Clin 2024). Supplementary denominators only; not used in EFR or avoidable-death computation.

### Section S7. Under-5 age-band sensitivity analysis

Under-5 sensitivity analysis. To test whether cross-country differences in 0–19 age structure confound the primary estimates, the entire EFR-frontier pipeline was recomputed on the single GBD under-5-years age group (age_id 1): country EFRs, super-region 10th-percentile frontiers and avoidable deaths = max(0, deaths − episodes × frontier EFR), for the same 204 countries and anchor years. Under-5 inputs are the official GBD 2023 age aggregates; for LRI in 2023 they are identical to the under-5 component used in the primary 0–19 pipeline (which sums the official <5 aggregate with the 5–9, 10–14 and 15–19 bands). Monte Carlo intervals for under-5 LRI avoidable deaths use the same 2,000-draw lognormal procedure as the primary analysis. In the under-5 band, composition effects from differing 0–19 age structures are removed by construction.

#### Table S7. Under-5 versus ages 0–19 estimates: deaths, episodes, episode-fatality ratios and avoidable deaths, by cause and year

| Cause | Year | Age band | Deaths | Episodes | EFR per 1,000 | Avoidable deaths | Avoidable share (%) |
|---|---|---|---|---|---|---|---|
| Lower respiratory infections | 1990 | Under 5 | 1,898,507 | 124,502,882 | 15.25 | 912,748 | 48.1 |
| Lower respiratory infections | 1990 | 0–19 | 2,035,714 | 166,066,423 | 12.26 | 923,895 | 45.4 |

| Cause | Year | Age band | Deaths | Episodes | EFR per 1,000 | Avoidable deaths | Avoidable share (%) |
|---|---|---|---|---|---|---|---|
| Lower respiratory infections | 2019 | Under 5 | 717,060 | 43,228,888 | 16.59 | 369,537 | 51.5 |
| Lower respiratory infections | 2019 | 0–19 | 816,491 | 71,525,886 | 11.42 | 391,968 | 48.0 |
| Lower respiratory infections | 2023 | Under 5 | 609,112 | 49,480,928 | 12.31 | 297,328 | 48.8 |
| Lower respiratory infections | 2023 | 0–19 | 711,228 | 80,998,085 | 8.78 | 333,803 | 46.9 |
| Upper respiratory infections | 1990 | Under 5 | 23,785 | 1,978,994,700 | 12.02 per M | 20,041 | 84.3 |
| Upper respiratory infections | 1990 | 0–19 | 27,128 | 5,108,264,596 | 5.31 per M | 22,196 | 81.8 |
| Upper respiratory infections | 2019 | Under 5 | 14,922 | 2,058,637,721 | 7.25 per M | 12,301 | 82.4 |
| Upper respiratory infections | 2019 | 0–19 | 17,290 | 5,560,945,204 | 3.11 per M | 13,982 | 80.9 |
| Upper respiratory infections | 2023 | Under 5 | 14,041 | 1,936,848,635 | 7.25 per M | 11,504 | 81.9 |
| Upper respiratory infections | 2023 | 0–19 | 16,377 | 5,611,244,034 | 2.92 per M | 13,273 | 81.0 |
| Whooping cough (pertussis) | 1990 | Under 5 | 233,528 | 25,851,914 | 9.03 | 91,067 | 39.0 |
| Whooping cough (pertussis) | 1990 | 0–19 | 277,403 | 30,194,376 | 9.19 | 104,378 | 37.6 |
| Whooping cough (pertussis) | 2023 | Under 5 | 95,523 | 15,562,668 | 6.14 | 35,669 | 37.3 |
| Whooping cough (pertussis) | 2023 | 0–19 | 112,839 | 18,469,250 | 6.11 | 43,958 | 39.0 |
| All three causes | 1990 | Under 5 | 2,155,820 | 2,129,349,495 | – | 1,023,856 | 47.5 |
| All three causes | 1990 | 0–19 | 2,340,244 | 5,304,525,395 | – | 1,050,468 | 44.9 |
| LRI + URI | 2019 | Under 5 | 731,982 | 2,101,866,609 | – | 381,838 | 52.2 |
| LRI + URI | 2019 | 0–19 | 833,781 | 5,632,471,090 | – | 405,950 | 48.7 |

| Cause | Year | Age band | Deaths | Episodes | EFR per 1,000 | Avoidable deaths | Avoidable share (%) |
| --- | --- | --- | --- | --- | --- | --- | --- |
| All three causes | 2023 | Under 5 | 718,676 | 2,001,892,230 | – | 344,500 | 47.9 |
| All three causes | 2023 | 0–19 | 840,444 | 5,710,711,369 | – | 391,034 | 46.5 |

EFR, episode-fatality ratio (deaths per incident episode); per M, per million episodes. Under-5 avoidable deaths use super-region 10th-percentile frontiers recomputed within the under-5 band; 0–19 rows repeat the primary estimates (main-text Tables 1, 2 and 4). Monte Carlo 95% UIs for under-5 avoidable LRI deaths: 800,523–1,087,007 (1990), 329,807–442,567 (2019) and 258,192– 365,192 (2023). Under-5 versus 0–19 country rankings, 2023: Spearman ρ = 0.98 (three causes) and 0.95 (LRI); identical top-5 countries in identical order (Nigeria, India, Niger, Democratic Republic of the Congo, Indonesia); top-10 share 62.2% versus 59.1%; Sub-Saharan Africa plus South Asia share 72.6% versus 73.1% (2023) and 42.2% versus 41.8% (1990); 2019–2023 change in avoidable LRI+URI deaths −19.1% versus −14.5%.
